# Predicting Alzheimer’s Disease Phenotypes With Aging Clocks: An Exploratory Analysis

**DOI:** 10.1101/2025.09.01.25334848

**Authors:** Cindy David Sarmento, Gabin Drouard, Toni T Saari, Aino Aaltonen, Aino Heikkinen, Teemu Palviainen, Sanna-Kaisa Herukka, Tarja Kokkola, Sari Kärkkäinen, FinnGen, Aarno Palotie, Valtteri Julkunen, Heiko Runz, Jaakko Kaprio, Miina Ollikainen, Eero Vuoksimaa

## Abstract

**Background:** The continuous aging of the world’s population urges the improvement of early diagnosis of age-related diseases, such as Alzheimer’s disease (AD). Blood proteomes can detect systemic and organ-specific disease-related changes and are accessible by minimally invasive blood draws.

**Methods:** We explored the potential of proteomic and epigenetic aging clocks as complementary biomarkers to established AD-related blood-based biomarkers (BBBs) and cognitive tests. Omics were generated from blood samples of 153 cognitively unimpaired individuals (average age 62 years). We investigated the associations of biological aging with BBBs and cognition, measured approximately nine years after the omics. We additionally tested whether dementia risk factors or genetic liability to them modulated these associations.

**Findings:** Accelerated systemic and brain-specific proteomic aging were linked with poorer cognitive functioning and higher plasma levels of neurofilament light chain. Interaction analysis showed that negative associations between proteomic aging and cognitive scores were stronger in individuals with lower genetic liability for type II diabetes. Altogether, proteomic clocks improved the explained variance in cognitive and biomarker measures by up to 18% compared with epigenetic clocks alone.

**Interpretation:** Our results support the potential of proteomes in detecting aging and AD-related phenotypes, particularly neurodegeneration and cognitive decline. However, co-morbidities may constitute confounding factors, highlighting the importance of further investigating proteomic aging in the context of AD using comprehensive approaches.

**Funding:** FIMM-EMBL International PhD Programme; Sigrid Jusélius Foundation; Academy of Finland; FinnGen.

## Introduction

Due to increasingly older populations, it has been estimated that over 150 million people will be affected by dementia globally in 2050, the equivalent of a 166% increase in just 30 years.^1^ Alzheimer’s disease (AD) is the most common cause of dementia. As the current practice often results in late diagnosis and a long lifetime spent with severe disability, AD represents a great burden to individuals, their families, and public health.^2^ Early diagnosis ensures opportunities for future financial and legal planning, and for early medical interventions to delay cognitive decline.^2,3^

AD is a highly complex and multifactorial neurodegenerative disease greatly dependent on genetics and potentially modifiable risk factors, with age recognized as the strongest risk factor.^4^ Aging-related molecular changes are typically interconnected, leading to physiological repercussions across several organs, and consequently, to the co-occurrence of several diseases.^5^ In recent years, advances in different omics have enabled the development of aging clocks. These tools predict biological age based on different molecular features and have successfully detected accelerated molecular aging associated with dementia.^6^ Some DNA methylation-based epigenetic clocks, such as DNAm PhenoAge and GrimAge, have shown significant associations between accelerated aging and poorer cognition as well as several AD hallmarks.^7,8^ However, findings in AD literature remain inconsistent, with multiple studies failing to find significant associations with AD phenotypes.^9^

Epigenetic clocks currently represent the most widely used approach to assess biological aging, but newly developed proteomic clocks hold great promise in AD research.^10,11^ The characteristic accumulation of beta-amyloid (Aβ) and phosphorylated tau (p-tau) aggregates in the brains of AD patients results from disruptions in protein homeostasis networks. This should be reflected in the blood proteomes and thus may be detectable by proteomic clocks.^12,13^ More recently, studies have introduced organ-specific proteomic clocks that evaluate the heterogeneity of organ aging and its impact on morbidity.^14–16^ Overall, these have provided further evidence that molecular aging processes are interconnected, as most fast-aging organs are associated with multiple diseases.^14–16^ Regarding dementia and AD, in addition to accelerated proteomic brain aging,^14,15^ fast-aging immune ^16^ and vascular ^14^ systems have also been associated with a higher incidence risk.

The Alzheimer’s Association workgroup’s revised guidelines from 2024 emphasized a more comprehensive approach to AD risk stratification to tackle the complexity added by common co-pathologies.^3^ Combining existing AD biomarkers and lifestyle and health data with molecular biomarkers of aging, including those captured by aging clocks, could provide better insights into an individual’s pathophysiology of AD before the occurrence of cognitive symptoms, which is essential to early diagnosis. However, to our knowledge, the utility of the recently developed proteomic aging clocks in predicting early cognitive impairment and state-of-the-art AD-related plasma biomarkers has not yet been evaluated in cognitively unimpaired individuals.

In this study, we leveraged an extensively phenotyped dataset of 153 individuals without a clinical diagnosis of AD to explore the utility of proteomic and epigenetic aging clocks in predicting AD-related phenotypes. We investigated the associations of aging clocks with blood-based biomarkers (BBBs) and cognitive screening tools. Additionally, we investigated common dementia risk factors, namely hypertension, hypercholesterolemia, and diabetes, as potential modifiers. We hypothesized that proteomic aging clocks, particularly those that are organ-specific and relevant to organs involved in AD, may better explain variability in BBBs and cognitive performance than epigenetic aging clocks.

### Research in context

#### Evidence before this study

On July 8, 2025, we conducted an advanced search in PubMed Central, which allows full-text searches without any language or publication date restrictions, using the terms ((proteomic OR epigenetic) AND aging clocks AND dementia). We identified a total of 1363 publications, the majority of which based on epigenetic data. Due to the characteristic disruption of proteostasis in AD, plasma proteomics has great potential as an early diagnostic tool. Although there is evidence correlating systemic, brain-, immune-, and artery-specific proteomic clocks with increased risk of dementia, their power in predicting early cognitive impairment and AD-related blood biomarkers remains underexplored at the population level.

#### Added value of this study

We examined associations of proteomic and epigenetic clocks with cognitive tests and state-of-the-art AD-related biomarkers in cognitively unimpaired individuals. We also integrated genetic and health questionnaire data to evaluate how common dementia risk factors modulated identified associations. To our knowledge, this was the first study leveraging such an extensively phenotyped sample of individuals without an AD diagnosis to evaluate proteomic clocks’ potential in early detection. Our analysis indicates that, collectively, proteomic aging clocks explain up to 18% of the variability in outcomes not accounted for by epigenetic clocks. But their performance was affected by genetic liability to diabetes.

#### Implications of all the available evidence

Our findings support the potential of plasma proteomics in the early detection of cognitive decline and neurodegeneration. Moreover, our results reinforce the importance of applying comprehensive approaches that include multiple omics, established AD blood biomarkers, cognitive testing, lifestyle, and health data, in AD risk assessment, to address the confounding effects of co-morbidities. Further research is needed in more diverse populations to evaluate the generalizability of proteomic clocks. Additionally, the influence of other dementia risk factors, medical interventions, and other possible confounding factors needs to be assessed.

## Methods

### Study design and participants

The population-based Older Finnish Twin Cohort (FTC) was established in 1974 with the recruitment of all Finnish same-sex twins born before 1958 residing in Finland.^17^ The baseline questionnaire had an approximate 90% participation rate, and follow-up data collections included questionnaires on health-related topics and in-person visits (Figure 1). Based on a 2011 questionnaire, twin pairs born between 1945 and 1957 discordant for blood pressure were identified and invited to participate in the Essential Hypertension Epigenetics (EH-Epi) study.^18^ From 2012 to 2015, participants visited the clinic, where blood samples were collected. From these, multi-omics data were generated, including the epigenetic and proteomic data used in this study. Additionally, genetic analyses were performed to compute polygenic risk scores (PRS) of dementia-related traits. Weight and height were also measured. In 2023, these individuals were invited to participate in the TWINGEN study.^19^

**Figure 1.**
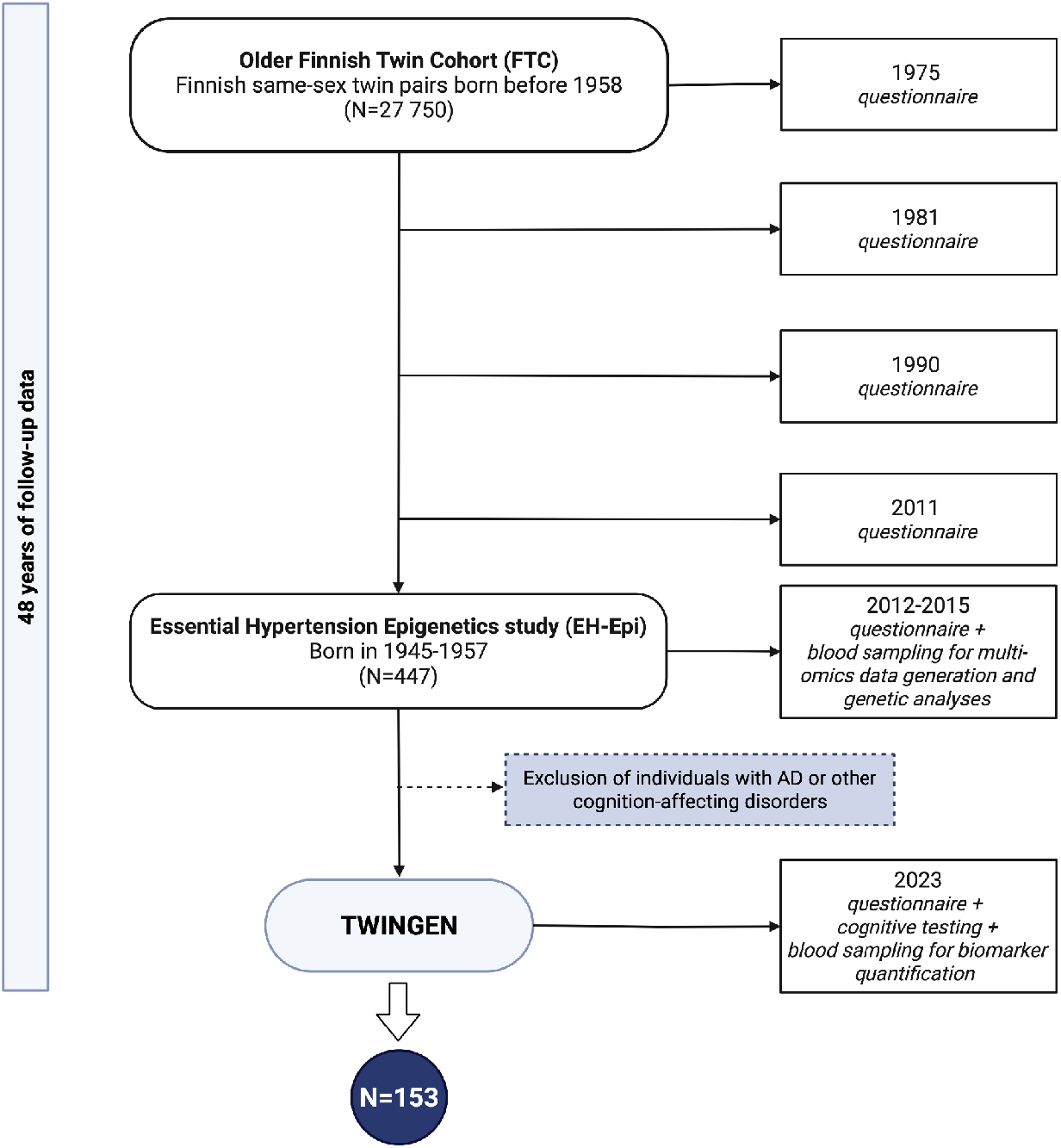
Flow chart of sample selection. Created in BioRender: https://BioRender.com/14pw0ri.

In TWINGEN, exclusion criteria were a diagnosis of AD or any other neurological or psychiatric disorder affecting cognition, based on health registry data and confirmation via telephone interview.^19^ Individuals were included in the study independently of their co-twin’s eligibility (n=153). The protocol included health questionnaires, telephone and computer-based cognitive testing, blood pressure measurements, and blood draws. These new blood samples were used to quantify AD-related plasma biomarkers.

### Outcomes: cognition and AD-related biomarkers

AD-related outcomes are summarized in Figure 2 and Table S1. Total scores from two dementia screening instruments were used as cognitive measures. These included a re-modified version of the Telephone Interview for Cognitive Status (TICS-m3)^20^ and the computer-administered tool cCOG. Both have been validated for early detection of cognitive impairment and AD against the gold-standard, in-person administered AD screening battery (in preprint ^20^). Additionally, Reaction Time (RT) was measured in the cCOG battery. This subtest was not included in the cCOG total score (Global Cognitive Score – GCS) but as a separate outcome.

**Figure 2.**
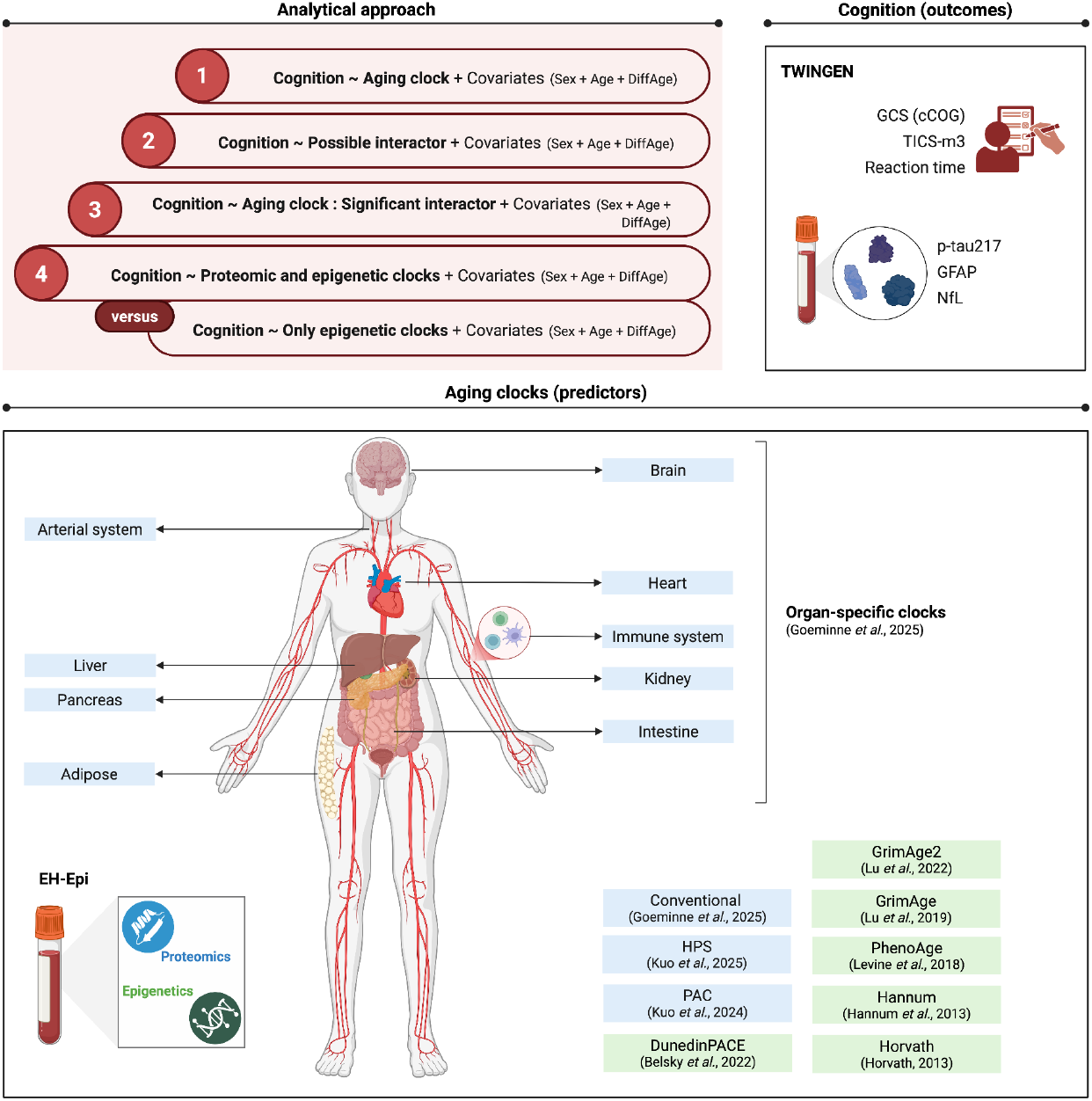
Analytical approach and variables. Abbreviations: GCS – global cognitive score for the cCOG test; TICS-m3 – Telephone Interview for Cognitive Status modified version 3; p-tau217 – phosphorylated tau 217; GFAP – glial fibrillary acidic protein; NfL – neurofilament light chain; HPS – Healthspan Proteomic Score; PAC – Proteomic Aging Clock. Created in BioRender: https://BioRender.com/1gxnv4y.

Among the BBBs considered in this study, plasma phosphorylated tau 217 (p-tau217) corresponds to an early-changing Core 1 biomarker, as defined by the Alzheimer’s Association workgroup in 2024.^3^ It is specific to AD neuropathological changes, reflecting both Aβ and tau pathologies. Neurofilament light chain (NfL) and glial fibrillary acidic proteins (GFAP), indicative of neurodegeneration and inflammation processes, respectively, are non-specific to AD, but suggested as possible prognostic markers.^3^ BBBs were quantified in TWINGEN blood samples using Simoa HD-X Analyzer (Quanterix, Billerica, Massachusetts, USA).^19^

### Predictors: epigenomic and proteomic aging clocks

DNA methylation and proteomic data were obtained from EH-Epi blood samples,^21^ collected eight to ten years before the TWINGEN study, and used to compute epigenetic and proteomic age estimates, respectively. DNA methylation (Illumina Infinium Human Methylation 450K Beadchip) and proteomic data (Olink Explore 3072) were processed for quality control as described by Drouard *et al*.^22^

Epigenetic age acceleration estimates were computed as in Drouard *et al*.^22^ by regressing chronological age onto the following epigenetic clock estimates: Horvath,^23^ Hannum,^24^ PhenoAge,^7^ GrimAge,^25^ GrimAge2,^26^ and DunedinPACE.^27^

We also employed recently developed second-generation systemic and organ-specific proteomic aging clocks, which are trained on mortality or morbidity instead of chronological age.^15^ These estimates correlate with chronological age and outperform other clocks in predicting risk of organ-specific diseases.^10,11,15^ System-wide proteomic clocks included Proteomic Aging Clock (PAC)^10^ and Healthspan Proteomic Score (HPS),^11^ which were estimated according to the developers’ instructions. In contrast with other clocks, which predict chronological age or mortality risk, HPS informs about the likelihood of remaining healthy, *i*.*e*., not developing any disease included in the healthspan definition (*e*.*g*., cancer, diabetes, heart failure, and dementia) nor dying within ten years from the baseline.^11^ Here, we used the estimate clocks. 1 – *HPS* to depict mortality hazard, in conformity with the other clocks.

We selected organ-specific clocks relevant to organs either directly (brain) or potentially involved in AD pathology, considering previously reported associations with different dementia types (arterial system, heart,^14^ immune system, and intestine^16^) or relation to major AD risk factors (diabetes – pancreas). The kidney-specific clock was included due to evidence that accelerated kidney aging has effects across multiple organs and is associated with diverse age-related diseases.^16^ Additionally, over-representation analysis of gene sets from each organ clock (Supplementary Materials) motivated the inclusion of the adipose- and liver-specific clocks due to significant enrichment in gene ontology terms and KEGG pathways relative to glucose transport and metabolism, type II diabetes (T2D), and acute inflammatory responses.

### Possible modifiers: dementia risk factors

We retrieved information on years of education, self-reported diabetes status (yes/no), measured glucose and cholesterol levels (elevated/normal) from available TWINGEN questionnaires. We also gathered measures of blood pressure taken in the context of the TWINGEN study and self-reported values when measures were not available. Additionally, we considered PRSs for AD, educational attainment (EA), T2D, fasting glucose (fg) and insulin (fi), coronary artery disease (CAD), low-density lipoprotein (LDL), and for systolic (SBP) and diastolic blood pressures (DBP).

### Statistical analyses

All statistical analyses were performed in R version 4.4.2.^28^ and can be summarized into three major steps (Figure 2):

(1) Association analysis between each cognition (cognitive tests and BBBs) and each biological age acceleration measure (epigenetic and proteomic clocks), using univariate modeling.
(2) Association analysis between cognition and possible interactors (*e*.*g*., diabetes status).
(3) Interaction analysis for each interactor and each aging clock significantly associated with cognition.
(4) Multivariate analysis with all clocks (full models) or only epigenetic clocks (reduced models) as predictors.

Generalized Estimating Equation (GEE) models were used for each step to account for the correlations within the data due to twins’ familial relatedness. For all models, covariates included sex assigned at birth obtained from a population registry, chronological age at first blood sampling, and time in years between first blood sampling and cognitive testing (DiffAge). All biological age estimates were normalized using rank-based inverse-normal transformation to unify the scales and correct distributional skewness. Additionally, continuous outcomes, predictors, and interactors were scaled. We reported T-values, 95% confidence intervals, and *p*-values (α = 0·05) adjusted for multiple testing with the Benjamini-Hochberg method in step (1), and nominal *p*-values in steps (2) and (3).

We performed interaction analyses to test the potential effect of genetic predisposition to AD and EA, as well as diabetes, hypertension, and hypercholesterolemia – common dementia risk factors – on associations between cognition and aging clocks. Possible interactors considered in the analyses were described in the previous section and summarized in Table S1. To correct for population structure, PRSs were regressed on the first ten genetic principal components and scaled in subsequent analyses. To interpret significant interactions, we stratified individuals into two PRS groups: below- and above-average. Then, we plotted biological age estimates against cognition estimates, fitting separate regression lines for each stratum.

Additionally, we conducted multivariate analysis to evaluate the collective value of proteomic clocks over epigenetic clocks in explaining variance in each outcome (Supplementary Materials). The conventional systemic second-generation proteomic clock was removed from this analysis due to high collinearity with other clocks (Table S7, Figure S11). Reduced models (epigenetic clocks only) and full models (epigenetic plus proteomic clocks) were adjusted for the same covariates (sex, age, and DiffAge). We reported the proportion of variance explained by the full models that is incrementally explained beyond the reduced models.

## Results

We investigated a total of 153 individuals, with an average age at blood sampling for omics data generation of approximately 62 years. Cognition and BBBs were measured on average nine years later (Table 1).

**Table 1.** General characteristics of the study sample. Age, weight, height, and BMI values correspond to values at the time of blood sampling in the EH-Epi study. DiffAge corresponds to the time gap between blood sampling and cognitive testing in the TWINGEN study.

|  | **Total** | **Female** | **Male** |
| --- | --- | --- | --- |
| **N** | 153 | 105 (69%) | 48 (31%) |
| **Zygosity** – MZ / DZ (N, %) | 92 (60%) / 61 (40%) | 67 / 38 | 25 / 23 |
| **Full pairs** – MZ / DZ (N) | 35 / 18 | 27 / 12 | 8 / 6 |
| **Age** – mean [SD] | 61·84 [3·59] | 61·91 [3·61] | 61·71 [3·58] |
| **DiffAge** – mean [SD] | 9·25 [0·70] | 9·24 [0·68] | 9·26 [0·75] |
| **Weight** – mean [SD] kg | 73·53 [14·01] | 69·36 [11·78] | 82·66 [14·29] |
| **Height** – mean [SD] cm | 166·45 [8·42] | 162·13 [5·22] | 175·90 [6·01] |
| **BMI** – mean [SD] | 26·47 [4·31] | 26·40 [4·52] | 26·63 [3·87] |
| **Education** – mean [SD] years | 12·53 [4·20] | 11·92 [3·95] | 13·85 [4·46] |

Abbreviations: MZ – monozygotic; DZ – dizygotic; BMI – body mass index; SD – standard deviation.

Associations between cognition (cognitive tests and BBBs) and aging clocks were reported in Figure 3.1 (additional results in Figure S1 and Table S2). Given the status of AD as an age-related disease, we expected accelerated aging detected by any of the clocks (higher estimates) to associate with lower cognitive performance in cCOG and TICS-m3 (negative coefficients), higher RT, and increased levels of BBBs (positive coefficients). Approximately 90% of significant associations detected involved proteomic clocks. Only one epigenetic clock, DunedinPACE, associated with an AD-related phenotype, specifically, decelerated pace of aging associated with higher levels of GFAP (T = -4·57 [95% CI -6·53 – -2·61]). Accelerated proteomic brain aging was associated with lower cognitive performance in both TICS-m3 (T = -3·04 [95% CI -5·00 – -1·08]) and cCOG (T = -3·29 [95% CI -5·25 – -1·33]), and higher plasma NfL levels (T = 4·14 [95% CI 2·18 – 6·10]). NfL showed the highest number of significant associations with biological aging, in particular, positive associations with multiple organ-specific and system-wide proteomic clocks. None of the clocks was significantly associated with either p-tau217 or RT.

**Figure 3.**
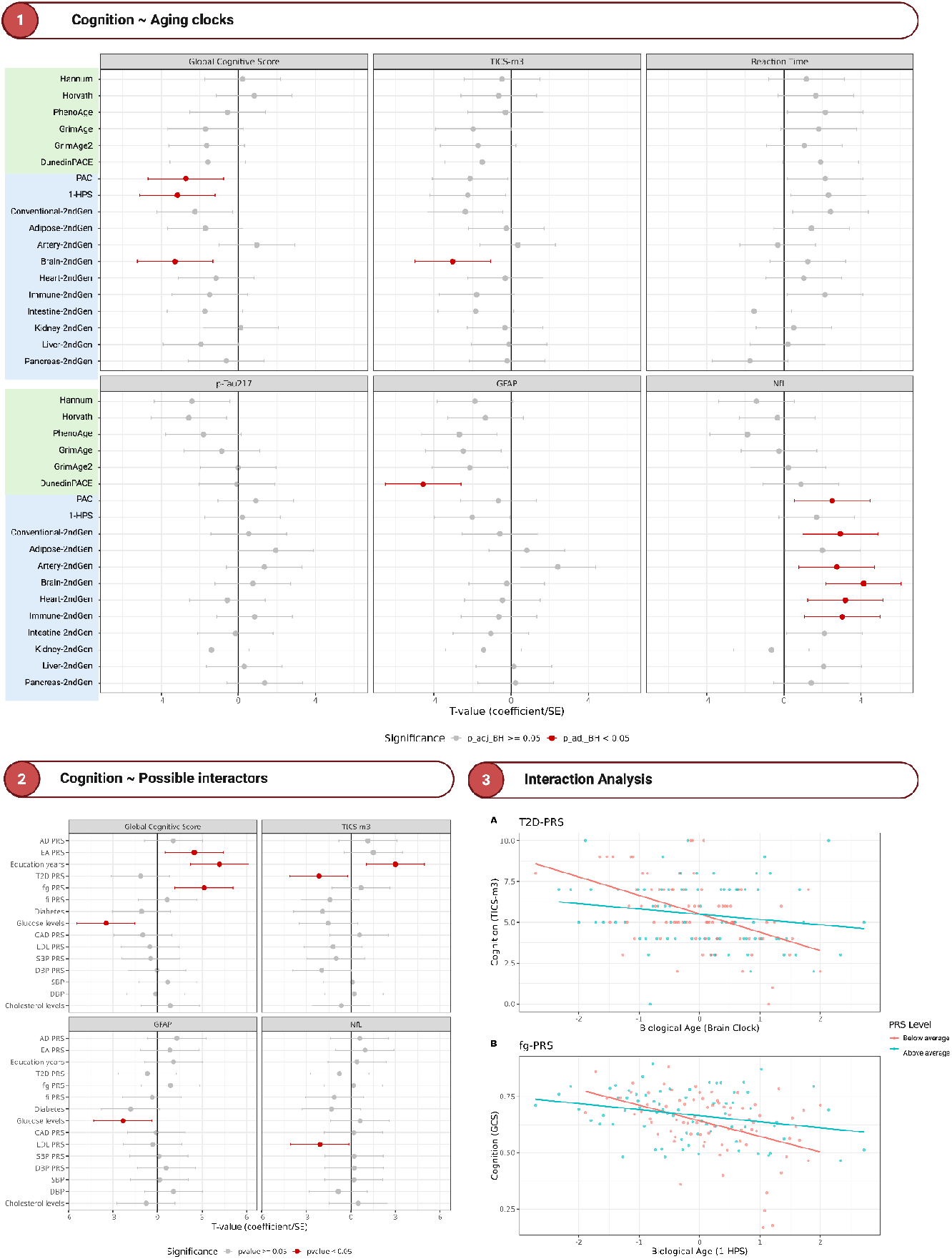
Association and interaction analyses’ results. (1) Associations of cognition measures and AD blood biomarkers with epigenetic and proteomic clocks. (2) Associations of cognition and AD blood biomarkers with nine PRSs, years of education, measured glucose, cholesterol, and blood pressure, and diabetes status. (3) Effect of type II diabetes and fasting glucose PRSs in cognition-biological age associations. Biological aging clocks are scaled. Statistically significant associations, defined as adjusted *p*-values below 0·05 in Panel 1 or nominal *p*-values below 0·05 in Panels 2 and 3, are highlighted in red. Created in BioRender: https://BioRender.com/vdzn1ga.

For all cognitive and biomarker measures that were significantly associated with an aging clock (GCS, TICS-m3 score, GFAP, and NfL), we further tested for possible interactors – indicators of diseases and AD risk factors, or genetic liability to them – and investigated if these modulated the cognition-clock associations (Figure 3.2, Table S3). Interaction analysis revealed two nominally significant interactions, suggesting T2D-PRS modulated the association between brain aging and TICS-m3 (*p* = 0·0186), and fg-PRS modulated the association between 1 – *HPS* and GCS (*p* = 0·0399; Table S4). More specifically, for individuals with above-average PRSs, the associations were weaker than in those with below-average PRSs (Figure 3.3).

Multivariate analysis revealed that including both proteomic and epigenetic clocks accounted for an additional 7 to 18% of the variance remaining after the epigenetic clocks and covariates alone (Table 2). Sensitivity analysis showed only minor decreases in model fit when dropping each proteomic clock (<5%; Table S8), failing to identify a single predictor with a particularly greater contribution to all outcomes.

**Table 2.** Multivariate analysis. Incremental R^2^ representing the proportion of residual variance in the reduced model (epigenetic clocks only) explained by adding proteomic clocks for each outcome.

| Outcome | Incremental $R^2$ |
| --- | --- |
| p-tau217 | 7·74% |
| Global Cognitive Score | 14·75% |
| TICS-m3 | 13·03% |
| Reaction Time | 18·19% |
| GFAP | 8·44% |
| NfL | 11·54% |

Abbreviations: p-tau217 – phosphorylated tau 217; TICS-m3 – Telephone Interview for Cognitive Status modified version 3; GFAP – glial fibrillary acidic protein; NfL – neurofilament light chain.

## Discussion

Our study leveraged an extensively phenotyped sample of cognitively unimpaired individuals to unveil potential associations between molecular aging and state-of-the-art markers of cognition and AD, as well as possible modifiers of these associations.

The results suggested that accelerated systemic and brain-specific proteomic aging are implicated in lower cognitive performance. Additionally, accelerated multi-organ aging was associated with higher plasma levels of NfL. These observations support the potential of plasma proteomics as auxiliary biomarkers of early cognitive decline and neurodegeneration. However, we did not observe significant associations between aging clocks and p-tau217, the only AD-specific outcome tested, suggesting clocks may be capturing general aging-related phenotypes rather than AD-specific changes. Notably, the brain-specific proteomic clock had moderate-to-high correlations with system-wide proteomic clocks (Pearson *r* = 0·69 – 0·76; Figure S11).

In addition to the brain-specific and systemic proteomic clocks, NfL was significantly associated with the artery-, heart-, and immune-specific clocks. Considering nominally significant *p*-values (Figure S1, Table S2), we retrieve additional associations with the adipose, intestine, and liver clocks. The variety in these associations may have multiple explanations. First, NfL is a non-AD-specific neurodegeneration biomarker that is increased in various diseases that involve neuronal loss.^3^ Therefore, individuals with multiple fast-aging organs may be more likely to show increased levels of NfL. Second, proteins used in clock modeling may be involved in processes or cellular components related to the nervous system or AD risk factors, driving AD pathogenesis and the associations with NfL. For example, enrichment analysis highlighted the aggrecan protein (*ACAN*) from the artery clock as a major component of perineuronal nets (PNNs; Figure S3). Prior reports have suggested that PNNs have protective functions against tau pathology: aggrecan binds to exogenous tau aggregates, trapping them in neurons’ extracellular space and preventing their internalization (see ^29^ for a review). Thus, changes in *ACAN* gene expression alter the composition of PNNs, disrupting their integrity and protective role.^29^ These changes may be captured by the artery clock, pointing to its potential in detecting AD-related pathologies.

Enrichment analysis also suggested possible confounding effects of AD risk factors on the associations. Proteins encoded by *BMP10* (bone morphogenetic protein 10), *MYL4* (myosin light chain 4), *TNNI3* (troponin I3), and *NPPB* (natriuretic peptide B), which are included in the heart-specific model, are involved in pathways regulating blood circulation and blood pressure (Figure S5). Additionally, proteins from the adipose clock indicated enrichment inKEGG’s T2D pathway. More specifically, adiponectin (*ADIPOQ*) and leptin (*LEP*) are concerned with several gene ontologies related to glucose metabolism and transport (Figure S2). This prompted us to perform an interaction analysis to clarify the influence of cardiovascular diseases and diabetes on the detected cognition-clock associations. Our findings indicated stronger negative associations between cognition and aging in individuals with below-average genetic liability for T2D and for high levels of fasting glucose. These results were consistent with a previous study that reported stronger associations between BBBs of AD, particularly p-tau217, and the levels of brain Aβ in those without diabetes or cardiovascular disorders.^30^ Altogether, these findings indicate that diabetes status may significantly affect the performance of proteomic clocks in predicting cognitive decline, which warrants further studies to validate.

Collectively, proteomic clocks explained up to 18% of the variance in cognition and AD-related biomarkers that was not captured by epigenetic clocks and covariates alone. These results support the hypothesis that plasma proteomes may detect additional aspects of AD pathophysiology that epigenetics do not. Yet, it is worth noting the imbalance in the number of epigenetic and proteomic clocks studied here – six and 12, respectively – as well as the differences in data types used to develop them and the magnitude of correlations between different proteomic clocks.

Our analyses included both first-generation clocks, trained on chronological age (Horvath and Hannum), and second-generation clocks, trained on either mortality or morbidity. Considering previous reports, we expected both epigenetic and proteomic mortality-based models to perform better than first-generation models in predicting risk of age-related diseases (*e*.*g*., ^7,15,25^). Additionally, DunedinPACE, developed based on longitudinal epigenetic data, constitutes a fundamentally distinct clock that could be classified as third-generation. It estimates not exactly biological age, but the pace at which aging and system decline occur.^27^ Notably, not only was DunedinPACE the sole epigenetic-based clock with a significant association with any outcome, it was the sole clock, overall, significantly associated with the non-AD-specific plasma biomarker of inflammation, GFAP. In the end, the performance of clocks in this context might depend not only on the omic layer from which they are derived but also on their methodological foundation.

Moreover, given the moderate-to-high correlations between some organ-specific and systemic proteomic clocks in our sample (Figure S11), it is not surprising that we observed collective associations between these clocks and several outcomes. Although the substantial correlations among proteomic clocks may inflate the number of reported associations, this overlap also suggests that the variance explained in the outcomes is likely shared across different proteomic clocks.

We acknowledge the limitations imposed on the generalizability of our results by the lack of racial and ethnic diversity in our dataset, which consisted exclusively of Finnish individuals with a homogeneous white European background. However, regarding educational background, our study sample provided more diversity than most, comprising individuals with poorer educational attainment as well as highly educated individuals. Additionally, we acknowledge that our study remains exploratory due to the small sample size that limits the statistical power of the analyses. We addressed this issue by using a remarkable variety of data types, encompassing genetics, epigenetics, and proteomics, as well as cognitive testing, health registry, and questionnaire data, enabling a more thorough investigation of AD risk.

To conclude, in our context of a small population-based sample of cognitively unimpaired individuals, the findings highlighted the added value of proteomic mortality-based clocks over epigenetic clocks in explaining variability in AD-related phenotypes and their potential as supplementary biomarkers of AD. Our study also underlined the importance of integrating multiple data modalities to address disease complexity. With this comprehensive approach, we showed that proteomic aging clocks may differentially capture variability in AD-related outcomes among individuals with above-average genetic risk for diabetes. Further research efforts to investigate the effect of other co-morbidities and risk factors, particularly those involving immunity and inflammation, should clarify the applicability of proteomics in risk assessment or clinical diagnosis. Longitudinal studies may also be beneficial in understanding how medical interventions targeting modifiable risk factors can modulate biological aging and cognitive functioning.

## Data Availability

TWINGEN data are stored at the THL Biobank for those participants who consented to data transfer to the biobank. Data are available to qualified applicants from academia and companies. For further details on the application process, see: https://thl.fi/en/research-and-development/thl-biobank/for-researchers/application-process.

## Author contributions

Conceptualization: CDS, GD, EV

Methodology: CDS, GD

Formal analysis: CDS

Investigation: CDS, GD, SH, TK, SJ, EV

Resources: VJ, JK, MO, AP, HR, EV, FinnGen

Data curation: CDS, GD, AH, TTS, AA

Writing – original draft: CDS, GD, EV

Writing – review and editing: all authors Visualization: CDS

Supervision: GD, EV

Funding acquisition: EV, FinnGen

## Ethics and consent statement

The TWINGEN protocol received ethical approval from the Coordinating Ethics Committee of the Hospital District of Helsinki and Uusimaa (HUS) (number 16831/2022), and the research plan was approved by THL Biobank (THLBB2022_83). All participants provided written informed consent before their participation and had the option to withdraw from the study at any point.

## Acknowledgments

We thank all participants in the Older Finnish Twin Cohort study who participated in the TWINGEN study, recruited through THL Biobank (study number THLBB2022_83).

C.D.S. was supported by the FIMM-EMBL International PhD Programme. E.V. was supported by the Sigrid Jusélius Foundation. J.K. was supported by the Academy of Finland Center of Excellence in Complex Disease Genetics (grant # 352792).

We want to acknowledge the participants and investigators of the FinnGen project, which funded the TWINGEN study. The FinnGen project is funded by two grants from Business Finland (HUS 4685/31/2016 and UH 4386/31/2016) and the following industry partners: AbbVie Inc., AstraZeneca UK Ltd, Biogen MA Inc., Bristol Myers Squibb Inc. (and Celgene Corporation & Celgene International II Sàrl), Genentech Inc., Merck Sharp & Dohme LCC, Pfizer Inc., GlaxoSmithKline Intellectual Property Development Ltd., Sanofi US Services Inc., Maze Therapeutics Inc., Johnson&Johnson Innovative Medicine Inc., Novartis AG, Boehringer Ingelheim International GmbH and Bayer AG. Following biobanks are acknowledged for delivering biobank samples to FinnGen: Auria Biobank (www.auria.fi/biopankki), THL Biobank (www.thl.fi/biobank), Helsinki Biobank (www.helsinginbiopankki.fi), Biobank Borealis of Northern Finland (https://www.ppshp.fi/Tutkimus-ja-opetus/Biopankki/Pages/Biobank-Borealis-briefly-in-English.aspx), Finnish Clinical Biobank Tampere (www.tays.fi/en-US/Research_and_development/Finnish_Clinical_Biobank_Tampere), Biobank of Eastern Finland (www.ita-suomenbiopankki.fi/en), Central Finland Biobank (www.ksshp.fi/fi-FI/Potilaalle/Biopankki), Finnish Red Cross Blood Service Biobank (www.veripalvelu.fi/verenluovutus/biopankkitoiminta), Terveystalo Biobank (www.terveystalo.com/fi/Yritystietoa/Terveystalo-Biopankki/Biopankki/) and Arctic Biobank (https://www.oulu.fi/en/university/faculties-and-units/faculty-medicine/northern-finland-birth-cohorts-and-arctic-biobank). All Finnish Biobanks are members of BBMRI.fi infrastructure (https://www.bbmri-eric.eu/national-nodes/finland/). Finnish Biobank Cooperative – FINBB (https://finbb.fi/) is the coordinator of BBMRI-ERIC operations in Finland. The Finnish biobank data can be accessed through the Fingenious® services (https://site.fingenious.fi/en/) managed by FINBB.

