## Supplementary Materials for "Predicting Alzheimer’s Disease Phenotypes With Aging Clocks: An Exploratory Analysis"

### Supplementary material

**Table of contents**

Extended methods---------------------------------------------------------------------------------------------------------------2

Table S1. Variables considered for association and interaction analyses. ---------------------------------------------2

Over-representation analysis (ORA)------------------------------------------------------------------------------------------3

Multivariate analysis -----------------------------------------------------------------------------------------------------------3

Extended results----------------------------------------------------------------------------------------------------------------4

Figure S1. Associations of cognition measures and AD blood biomarkers with epigenetic and proteomic clocks. -----------------------------------------------------------------------------------------------------------------------------4

Table S2. Associations between aging clocks and AD-related phenotypes in univariate analysis.------------------5

Table S3. Associations between measures representative of dementia risk factors (modifiers) and AD-related phenotypes.--------------------------------------------------------------------------------------------------------------------9

Table S4. Interactions between modifiers and aging clocks significantly associated with AD-related outcomes. ---------------------------------------------------------------------------------------------------------------------------- 11

Table S5. Top 30 GO categories significantly enriched in each organ-specific proteomic clock.------------------ 12

Table S6. Top 30 KEGG pathways significantly enriched in each organ-specific proteomic clock. ------------- 22

Figure S2. Top 30 GO categories and KEGG pathways significantly enriched in the adipose clock. ------------ 24

Figure S3. Top 30 GO categories and KEGG pathways significantly enriched in the artery clock. -------------- 25

Figure S4. Top 30 GO categories and KEGG pathways significantly enriched in the brain clock. ---------------- 26

Figure S5. Top 30 GO categories and KEGG pathways significantly enriched in the heart clock. ---------------- 27

Figure S6. Top 30 GO categories and KEGG pathways significantly enriched in the immune clock. ----------- 28

Figure S7. Top 30 GO categories and KEGG pathways significantly enriched in the intestine clock. ----------- 29

Figure S8. Top 30 GO categories and KEGG pathways significantly enriched in the kidney clock. ------------- 30

Figure S9. Top 30 GO categories and KEGG pathways significantly enriched in the liver clock.----------------- 31

Figure S10. Top 30 GO categories and KEGG pathways significantly enriched in the pancreas clock. ---------- 32

Table S7. Variance inflation factors for each predictor (clock) considered for multivariate analysis.------------ 33

Figure S11. Pearson's correlations between each proteomic and epigenetic aging clock. -------------------------- 34

Table S8. Decrease in pseudo-$R^2$ in multivariate submodels.------------------------------------------------------------ 35

References---------------------------------------------------------------------------------------------------------------------- 36

### Extended methods

**Table S1. Variables considered for association and interaction analyses.**

| Class | Variables | Reference (PMID) | Origin of data |
|---|---|---|---|
| **Outcomes** | cCOG Global Cognitive Score (GCS) | [1] | TWINGEN (2023) |
| | TICS-m3 | | |
| | Reaction Time | [2] 38866570 | |
| | p-tau217 | [3] 40352683 | |
| | GFAP | [2] 38866570 | |
| | NfL | | |
| **Predictors** | Hannum | [4] 23177740 | EH-Epi blood samples (2012-2015) |
| | Horvath | [5] 24138928 | |
| | PhenoAge | [6] 29676998 | |
| | GrimAge | [7] 30669119 | |
| | GrimAge2 | [8] 36516495 | |
| | DunedinPACE | [9] 35029144 | |
| | PAC | [10] 38747160 | |
| | 1 - HPS | [11] 38978645 | |
| | Conventional | [12] 39488213 | |
| | Adipose | | |
| | Artery | | |
| | Brain | | |
| | Heart | | |
| | Immune | | |
| | Intestine | | |
| | Kidney | | |
| | Liver | | |
| | Pancreas | | |
| **Covariates** | Sex | .. | .. |
| | Age | | |
| | Time gap between sampling (DiffAge) | | |
| **Interactors** | AD PRS | 24162737 | EH-Epi blood samples (2012-2015) |
| | EA PRS | 30038396 | |
| | T2D PRS | 30297969 | |

| Class | Variables | Reference (PMID) | Origin of data |
|---|---|---|---|
| | fg PRS | 34059833 | |
| | fi PRS | | |
| | CAD PRS | 26343387 | |
| | LDL PRS | 34887591 | |
| | SBP PRS (4080_raw) | UKBB http://www.nealelab.is/uk-biobank/ | |
| | DBP PRS (4079_raw) | | |
| | Education in years | .. | TWINGEN (2023) |
| | Diabetes status | | |
| | Glucose levels | | |
| | Cholesterol levels | | |
| | Blood pressure levels | | |

Abbreviations: PMID – PubMed Identifier; TICS-m3 – Telephone Interview for Cognitive Status modified version 3; p-tau217 – phosphorylated tau 217; GFAP – glial fibrillary acidic protein; NfL – neurofilament light chain; PAC – Proteomic Aging Clock; HPS – Healthspan Proteomic Score; PRS – polygenic risk score; AD – Alzheimer's Disease; EA – educational attainment; T2D – type II diabetes; fg – fasting glucose; fi – fasting insulin; CAD – coronary artery disease; LDL – low-density lipoprotein; SBP – systolic blood pressure; DBP – diastolic blood pressure; UKBB – UK Biobank.

**Over-representation analysis (ORA)**

To determine whether certain biological functions and molecular interaction pathways were enriched in the list of proteins from each organ-specific clock, we performed over-representation analysis (ORA). We used the tools *enrichGO* and *enrichKEGG* from the *clusterProfiler* R package v.4.14.6 [13], which only require gene identifiers to compute enrichment of all GO (Gene Ontology) and KEGG (Kyoto Encyclopedia of Genes and Genomes) categories. Protein-coding gene names were converted to Entrez IDs using the tool *bitr,* also implemented in the *clusterProfiler* package. We set the *p*-value cutoff to 0·05 and chose the Benjamini-Hochberg method for false-discovery rate control. Results are presented in Tables S5-S6 and Figures S2-S10.

**Multivariate analysis**

To assess the additional value of proteomic clocks over epigenetic clocks in predicting cognitive function, we performed multivariate analysis. Collinearity among aging clocks was estimated by computing variance inflation factors (VIF) on linear regression models for each outcome (Table S7) and Pearson's correlations (Figure S11). Most clocks had low to moderate collinearity (VIF < 10). Only the second-generation systemic proteomic clock (Conventional_2ndGen) exhibited strong collinearity (VIF > 12) concurrent with very high correlation with two other proteomic clocks, $1 - HPS$ (0·86) and PAC (0·92)*,* and was thus removed from multivariate models.
We ran full multivariate models for each outcome, including all proteomic and epigenetic clocks as predictors (except the Conventional_2ndGen) and sex, age, and DiffAge as covariates. Reduced multivariate models included only epigenetic clocks as predictors, adjusted for the same covariates. To measure the improvement in fit between the full and reduced models, we computed Zheng's pseudo-$R^2$, an extension of $R^2$ statistics for Generalized Estimating Equation models proposed by Zheng *et al.* [14]. Additionally, we performed sensitivity analysis to assess the importance of each proteomic clock in improving model fit. For that, we ran submodels, dropping each predictor at a time, and computed the difference between the full model's $R^2$ and the submodels' $R^2$ (Table S8).

### Extended results

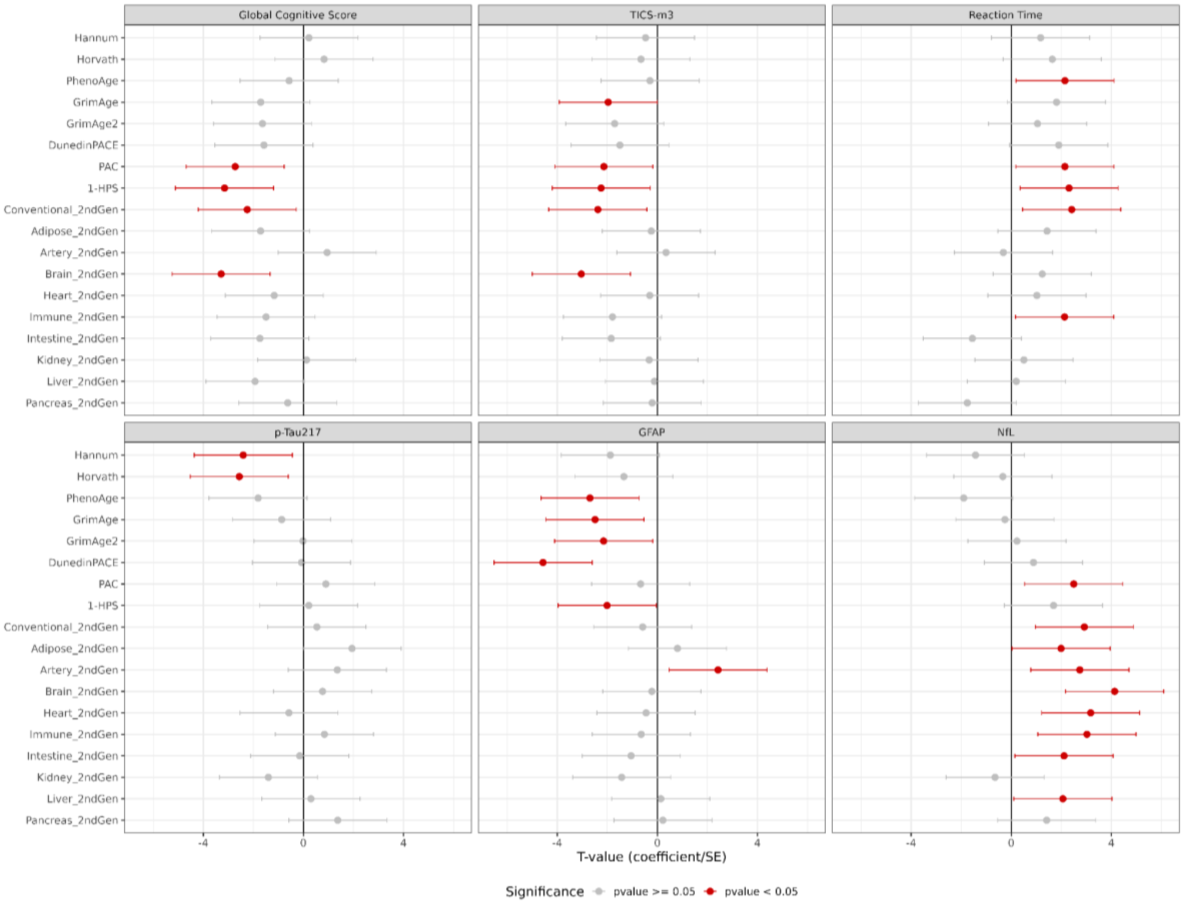


**Figure S1. Associations of cognition measures and AD blood biomarkers with epigenetic and proteomic clocks.**

All associations with nominal *p*-values below 0·05 are highlighted in red. Biological aging clocks are scaled.

**Table S2. Associations between aging clocks and AD-related phenotypes in univariate analysis.**

| Outcome | Predictor | β-coefficient | SE | 95% CI Lower | 95% CI Upper | T-value | *p*-value | Adjusted *p*-value (BH) | Adjusted *p*-value (Bonferroni) |
|---|---|---|---|---|---|---|---|---|---|
| **GCS** | Hannum | 0·0196 | 0·0888 | -1·7391 | 2·1809 | 0·2209 | 0·8251 | 0·8737 | 1·0000 |
| | Horvath | 0·0716 | 0·0867 | -1·1343 | 2·7857 | 0·8257 | 0·4090 | 0·5258 | 1·0000 |
| | PhenoAge | -0·0466 | 0·0817 | -2·5310 | 1·3890 | -0·5710 | 0·5680 | 0·6390 | 1·0000 |
| | GrimAge | -0·1641 | 0·0962 | -3·6660 | 0·2540 | -1·7060 | 0·0880 | 0·1980 | 1·0000 |
| | GrimAge2 | -0·1293 | 0·0790 | -3·5963 | 0·3237 | -1·6363 | 0·1018 | 0·2035 | 1·0000 |
| | DunedinPACE | -0·1135 | 0·0719 | -3·5387 | 0·3813 | -1·5787 | 0·1144 | 0·2059 | 1·0000 |
| | PAC | -0·2589 | 0·0950 | -4·6859 | -0·7659 | -2·7259 | 0·0064 | 0·0385 | 0·1154 |
| | 1-HPS | -0·2778 | 0·0881 | -5·1134 | -1·1934 | -3·1534 | 0·0016 | 0·0145 | 0·0291 |
| | Conventional_2ndGen | -0·2213 | 0·0985 | -4·2066 | -0·2866 | -2·2466 | 0·0247 | 0·1110 | 0·4440 |
| | Adipose_2ndGen | -0·1766 | 0·1031 | -3·6720 | 0·2480 | -1·7120 | 0·0869 | 0·1980 | 1·0000 |
| | Artery_2ndGen | 0·0864 | 0·0911 | -1·0118 | 2·9082 | 0·9482 | 0·3431 | 0·4750 | 1·0000 |
| | Brain_2ndGen | -0·2895 | 0·0881 | -5·2474 | -1·3274 | -3·2874 | 0·0010 | 0·0145 | 0·0182 |
| | Heart_2ndGen | -0·0982 | 0·0843 | -3·1255 | 0·7945 | -1·1655 | 0·2438 | 0·3657 | 1·0000 |
| | Immune_2ndGen | -0·1345 | 0·0900 | -3·4544 | 0·4656 | -1·4944 | 0·1351 | 0·2210 | 1·0000 |
| | Intestine_2ndGen | -0·1279 | 0·0735 | -3·6996 | 0·2204 | -1·7396 | 0·0819 | 0·1980 | 1·0000 |
| | Kidney_2ndGen | 0·0100 | 0·0800 | -1·8344 | 2·0856 | 0·1256 | 0·9000 | 0·9000 | 1·0000 |
| | Liver_2ndGen | -0·1752 | 0·0907 | -3·8927 | 0·0273 | -1·9327 | 0·0533 | 0·1918 | 0·9588 |
| | Pancreas_2ndGen | -0·0554 | 0·0882 | -2·5877 | 1·3323 | -0·6277 | 0·5302 | 0·6362 | 1·0000 |
| **TICS-m3** | Hannum | -0·0471 | 0·0999 | -2·4314 | 1·4886 | -0·4714 | 0·6374 | 0·8880 | 1·0000 |
| | Horvath | -0·0691 | 0·1057 | -2·6135 | 1·3065 | -0·6535 | 0·5134 | 0·8880 | 1·0000 |
| | PhenoAge | -0·0249 | 0·0851 | -2·2526 | 1·6674 | -0·2926 | 0·7699 | 0·8880 | 1·0000 |
| | GrimAge | -0·1719 | 0·0875 | -3·9243 | -0·0043 | -1·9643 | 0·0495 | 0·1782 | 0·8908 |
| | GrimAge2 | -0·1354 | 0·0794 | -3·6648 | 0·2552 | -1·7048 | 0·0882 | 0·1985 | 1·0000 |
| | DunedinPACE | -0·1140 | 0·0762 | -3·4561 | 0·4639 | -1·4961 | 0·1346 | 0·2693 | 1·0000 |
| | PAC | -0·1795 | 0·0841 | -4·0956 | -0·1756 | -2·1356 | 0·0327 | 0·1472 | 0·5888 |
| | 1-HPS | -0·1889 | 0·0841 | -4·2055 | -0·2855 | -2·2455 | 0·0247 | 0·1472 | 0·4452 |
| | Conventional_2ndGen | -0·2016 | 0·0849 | -4·3361 | -0·4161 | -2·3761 | 0·0175 | 0·1472 | 0·3149 |
| | Adipose_2ndGen | -0·0177 | 0·0732 | -2·2018 | 1·7182 | -0·2418 | 0·8090 | 0·8880 | 1·0000 |
| | Artery_2ndGen | 0·0261 | 0·0752 | -1·6129 | 2·3071 | 0·3471 | 0·7285 | 0·8880 | 1·0000 |
| | Brain_2ndGen | -0·2611 | 0·0859 | -4·9998 | -1·0798 | -3·0398 | 0·0024 | 0·0426 | 0·0426 |
| | Heart_2ndGen | -0·0291 | 0·0956 | -2·2643 | 1·6557 | -0·3043 | 0·7609 | 0·8880 | 1·0000 |
| | Immune_2ndGen | -0·1350 | 0·0753 | -3·7519 | 0·1681 | -1·7919 | 0·0731 | 0·1881 | 1·0000 |

| | | | | | | | | |
|---|---|---|---|---|---|---|---|---|
| | Intestine_2ndGen | -0·1506 | 0·0818 | -3·8017 | 0·1183 | -1·8417 | 0·0655 | 0·1881 | 1·0000 |
| | Kidney_2ndGen | -0·0273 | 0·0836 | -2·2866 | 1·6334 | -0·3266 | 0·7439 | 0·8880 | 1·0000 |
| | Liver_2ndGen | -0·0087 | 0·0756 | -2·0751 | 1·8449 | -0·1151 | 0·9084 | 0·9084 | 1·0000 |
| | Pancreas_2ndGen | -0·0179 | 0·0878 | -2·1636 | 1·7564 | -0·2036 | 0·8387 | 0·8880 | 1·0000 |
| **RT** | Hannum | 0·1161 | 0·0989 | -0·7861 | 3·1339 | 1·1739 | 0·2404 | 0·3329 | 1·0000 |
| | Horvath | 0·1808 | 0·1100 | -0·3164 | 3·6036 | 1·6436 | 0·1003 | 0·2005 | 1·0000 |
| | PhenoAge | 0·1838 | 0·0856 | 0·1877 | 4·1077 | 2·1477 | 0·0317 | 0·1189 | 0·5713 |
| | GrimAge | 0·1925 | 0·1063 | -0·1484 | 3·7716 | 1·8116 | 0·0700 | 0·1783 | 1·0000 |
| | GrimAge2 | 0·0890 | 0·0848 | -0·9099 | 3·0101 | 1·0501 | 0·2937 | 0·3655 | 1·0000 |
| | DunedinPACE | 0·1608 | 0·0847 | -0·0605 | 3·8595 | 1·8995 | 0·0575 | 0·1725 | 1·0000 |
| | PAC | 0·2097 | 0·0978 | 0·1842 | 4·1042 | 2·1442 | 0·0320 | 0·1189 | 0·5763 |
| | 1-HPS | 0·1971 | 0·0852 | 0·3519 | 4·2719 | 2·3119 | 0·0208 | 0·1189 | 0·3741 |
| | Conventional_2ndGen | 0·2217 | 0·0917 | 0·4586 | 4·3786 | 2·4186 | 0·0156 | 0·1189 | 0·2804 |
| | Adipose_2ndGen | 0·1261 | 0·0881 | -0·5288 | 3·3912 | 1·4312 | 0·1524 | 0·2493 | 1·0000 |
| | Artery_2ndGen | -0·0275 | 0·0878 | -2·2737 | 1·6463 | -0·3137 | 0·7538 | 0·7981 | 1·0000 |
| | Brain_2ndGen | 0·1129 | 0·0910 | -0·7197 | 3·2003 | 1·2403 | 0·2149 | 0·3223 | 1·0000 |
| | Heart_2ndGen | 0·1170 | 0·1140 | -0·9333 | 2·9867 | 1·0267 | 0·3046 | 0·3655 | 1·0000 |
| | Immune_2ndGen | 0·1775 | 0·0833 | 0·1716 | 4·0916 | 2·1316 | 0·0330 | 0·1189 | 0·5947 |
| | Intestine_2ndGen | -0·1494 | 0·0962 | -3·5132 | 0·4068 | -1·5532 | 0·1204 | 0·2167 | 1·0000 |
| | Kidney_2ndGen | 0·0456 | 0·0899 | -1·4531 | 2·4669 | 0·5069 | 0·6123 | 0·6888 | 1·0000 |
| | Liver_2ndGen | 0·0188 | 0·0941 | -1·7604 | 2·1596 | 0·1996 | 0·8418 | 0·8418 | 1·0000 |
| | Pancreas_2ndGen | -0·1526 | 0·0870 | -3·7152 | 0·2048 | -1·7552 | 0·0792 | 0·1783 | 1·0000 |
| **p-Tau217** | Hannum | -0·2233 | 0·0929 | -4·3653 | -0·4453 | -2·4053 | 0·0162 | 0·1454 | 0·2909 |
| | Horvath | -0·2052 | 0·0801 | -4·5218 | -0·6018 | -2·5618 | 0·0104 | 0·1454 | 0·1874 |
| | PhenoAge | -0·1653 | 0·0912 | -3·7722 | 0·1478 | -1·8122 | 0·0700 | 0·3148 | 1·0000 |
| | GrimAge | -0·0643 | 0·0741 | -2·8283 | 1·0917 | -0·8683 | 0·3852 | 0·7211 | 1·0000 |
| | GrimAge2 | -0·0011 | 0·0675 | -1·9765 | 1·9435 | -0·0165 | 0·9868 | 0·9868 | 1·0000 |
| | DunedinPACE | -0·0052 | 0·0704 | -2·0341 | 1·8859 | -0·0741 | 0·9410 | 0·9868 | 1·0000 |
| | PAC | 0·0719 | 0·0801 | -1·0627 | 2·8573 | 0·8973 | 0·3696 | 0·7211 | 1·0000 |
| | 1-HPS | 0·0143 | 0·0672 | -1·7464 | 2·1736 | 0·2136 | 0·8309 | 0·9868 | 1·0000 |
| | Conventional_2ndGen | 0·0415 | 0·0772 | -1·4222 | 2·4978 | 0·5378 | 0·5907 | 0·8179 | 1·0000 |
| | Adipose_2ndGen | 0·1285 | 0·0662 | -0·0187 | 3·9013 | 1·9413 | 0·0522 | 0·3133 | 0·9400 |
| | Artery_2ndGen | 0·1074 | 0·0793 | -0·6053 | 3·3147 | 1·3547 | 0·1755 | 0·4513 | 1·0000 |
| | Brain_2ndGen | 0·0657 | 0·0860 | -1·1960 | 2·7240 | 0·7640 | 0·4449 | 0·7280 | 1·0000 |

| | | | | | | | | | |
|---|---|---|---|---|---|---|---|---|---|
| | Heart_2ndGen | -0·0574 | 0·0991 | -2·5394 | 1·3806 | -0·5794 | 0·5623 | 0·8179 | 1·0000 |
| | Immune_2ndGen | 0·0463 | 0·0551 | -1·1195 | 2·8005 | 0·8405 | 0·4006 | 0·7211 | 1·0000 |
| | Intestine_2ndGen | -0·0093 | 0·0621 | -2·1095 | 1·8105 | -0·1495 | 0·8812 | 0·9868 | 1·0000 |
| | Kidney_2ndGen | -0·0937 | 0·0669 | -3·3597 | 0·5603 | -1·3997 | 0·1616 | 0·4513 | 1·0000 |
| | Liver_2ndGen | 0·0196 | 0·0649 | -1·6587 | 2·2613 | 0·3013 | 0·7632 | 0·9812 | 1·0000 |
| | Pancreas_2ndGen | 0·1024 | 0·0746 | -0·5877 | 3·3323 | 1·3723 | 0·1700 | 0·4513 | 1·0000 |
| **GFAP** | Hannum | -0·1587 | 0·0845 | -3·8387 | 0·0813 | -1·8787 | 0·0603 | 0·1550 | 1·0000 |
| | Horvath | -0·1084 | 0·0811 | -3·2961 | 0·6239 | -1·3361 | 0·1815 | 0·3630 | 1·0000 |
| | PhenoAge | -0·2182 | 0·0810 | -4·6528 | -0·7328 | -2·6928 | 0·0071 | 0·0638 | 0·1275 |
| | GrimAge | -0·2124 | 0·0854 | -4·4479 | -0·5279 | -2·4879 | 0·0128 | 0·0688 | 0·2313 |
| | GrimAge2 | -0·1784 | 0·0830 | -4·1081 | -0·1881 | -2·1481 | 0·0317 | 0·1141 | 0·5707 |
| | DunedinPACE | -0·2949 | 0·0646 | -6·5276 | -2·6076 | -4·5676 | 0·000005 | 0·0001 | 0·0001 |
| | PAC | -0·0527 | 0·0788 | -2·6292 | 1·2908 | -0·6692 | 0·5034 | 0·7184 | 1·0000 |
| | 1-HPS | -0·1312 | 0·0653 | -3·9686 | -0·0486 | -2·0086 | 0·0446 | 0·1337 | 0·8024 |
| | Conventional_2ndGen | -0·0433 | 0·0741 | -2·5447 | 1·3753 | -0·5847 | 0·5587 | 0·7184 | 1·0000 |
| | Adipose_2ndGen | 0·0617 | 0·0767 | -1·1550 | 2·7650 | 0·8050 | 0·4208 | 0·6886 | 1·0000 |
| | Artery_2ndGen | 0·1833 | 0·0756 | 0·4654 | 4·3854 | 2·4254 | 0·0153 | 0·0688 | 0·2753 |
| | Brain_2ndGen | -0·0183 | 0·0835 | -2·1792 | 1·7408 | -0·2192 | 0·8265 | 0·8751 | 1·0000 |
| | Heart_2ndGen | -0·0276 | 0·0612 | -2·4104 | 1·5096 | -0·4504 | 0·6524 | 0·7829 | 1·0000 |
| | Immune_2ndGen | -0·0448 | 0·0696 | -2·6043 | 1·3157 | -0·6443 | 0·5194 | 0·7184 | 1·0000 |
| | Intestine_2ndGen | -0·0767 | 0·0731 | -3·0086 | 0·9114 | -1·0486 | 0·2944 | 0·5298 | 1·0000 |
| | Kidney_2ndGen | -0·1164 | 0·0817 | -3·3843 | 0·5357 | -1·4243 | 0·1544 | 0·3473 | 1·0000 |
| | Liver_2ndGen | 0·0103 | 0·0733 | -1·8192 | 2·1008 | 0·1408 | 0·8880 | 0·8880 | 1·0000 |
| | Pancreas_2ndGen | 0·0168 | 0·0754 | -1·7369 | 2·1831 | 0·2231 | 0·8235 | 0·8751 | 1·0000 |
| **NfL** | Hannum | -0·0809 | 0·0567 | -3·3860 | 0·5340 | -1·4260 | 0·1539 | 0·2165 | 1·0000 |
| | Horvath | -0·0170 | 0·0506 | -2·2956 | 1·6244 | -0·3356 | 0·7372 | 0·8185 | 1·0000 |
| | PhenoAge | -0·1471 | 0·0775 | -3·8576 | 0·0624 | -1·8976 | 0·0578 | 0·1040 | 1·0000 |
| | GrimAge | -0·0159 | 0·0634 | -2·2116 | 1·7084 | -0·2516 | 0·8013 | 0·8185 | 1·0000 |
| | GrimAge2 | 0·0124 | 0·0541 | -1·7305 | 2·1895 | 0·2295 | 0·8185 | 0·8185 | 1·0000 |
| | DunedinPACE | 0·0440 | 0·0495 | -1·0709 | 2·8491 | 0·8891 | 0·3740 | 0·4808 | 1·0000 |
| | PAC | 0·1499 | 0·0600 | 0·5381 | 4·4581 | 2·4981 | 0·0125 | 0·0375 | 0·2247 |
| | 1-HPS | 0·0893 | 0·0529 | -0·2699 | 3·6501 | 1·6901 | 0·0910 | 0·1489 | 1·0000 |
| | Conventional_2ndGen | 0·2454 | 0·0840 | 0·9623 | 4·8823 | 2·9223 | 0·0035 | 0·0156 | 0·0625 |
| | Adipose_2ndGen | 0·0949 | 0·0476 | 0·0328 | 3·9528 | 1·9928 | 0·0463 | 0·0926 | 0·8331 |

| | | | | | | | | |
|---|---|---|---|---|---|---|---|---|
| | Artery_2ndGen | 0·2621 | 0·0956 | 0·7823 | 4·7023 | 2·7423 | 0·0061 | 0·0220 | 0·1098 |
| | Brain_2ndGen | 0·2234 | 0·0540 | 2·1751 | 6·0951 | 4·1351 | 0·00004 | 0·0006 | 0·0006 |
| | Heart_2ndGen | 0·1831 | 0·0577 | 1·2152 | 5·1352 | 3·1752 | 0·0015 | 0·0135 | 0·0269 |
| | Immune_2ndGen | 0·1538 | 0·0508 | 1·0660 | 4·9860 | 3·0260 | 0·0025 | 0·0149 | 0·0446 |
| | Intestine_2ndGen | 0·1063 | 0·0503 | 0·1518 | 4·0718 | 2·1118 | 0·0347 | 0·0872 | 0·6247 |
| | Kidney_2ndGen | -0·0602 | 0·0931 | -2·6063 | 1·3137 | -0·6463 | 0·5181 | 0·6217 | 1·0000 |
| | Liver_2ndGen | 0·1456 | 0·0704 | 0·1065 | 4·0265 | 2·0665 | 0·0388 | 0·0872 | 0·6980 |
| | Pancreas_2ndGen | 0·0860 | 0·0607 | -0·5425 | 3·3775 | 1·4175 | 0·1563 | 0·2165 | 1·0000 |

Abbreviations: SE – standard error; BH – Benjamini-Hochberg method for multiple testing correction; CI – confidence interval; GCS – Global Cognitive Score; TICS-m3 – Telephone Interview for Cognitive Status modified version 3; RT – Reaction Time; p-tau217 – phosphorylated tau 217; GFAP – glial fibrillary acidic protein; NfL – neurofilament light chain; PAC – Proteomic Aging Clock; HPS – Healthspan Proteomic Score.

**Table S3. Associations between measures representative of dementia risk factors (modifiers) and AD-related phenotypes.**

| Outcome | Predictor | β-coefficient | SE | 95% CI Lower | 95% CI Upper | T-value | *p*-value |
|---|---|---|---|---|---|---|---|
| **GCS** | AD PRS | 0·0845 | 0·0795 | -0·8963 | 3·0237 | 1·0637 | 0·2875 |
| | EA PRS | 0·1892 | 0·0764 | 0·5162 | 4·4362 | 2·4762 | 0·0133 |
| | Education years | 0·3016 | 0·0722 | 2·2157 | 6·1357 | 4·1757 | 0·00003 |
| | T2D PRS | -0·0836 | 0·0730 | -3·1043 | 0·8157 | -1·1443 | 0·2525 |
| | fg PRS | 0·2619 | 0·0835 | 1·1761 | 5·0961 | 3·1361 | 0·0017 |
| | fi PRS | 0·0647 | 0·0983 | -1·3023 | 2·6177 | 0·6577 | 0·5107 |
| | Diabetes | -0·2612 | 0·2425 | -3·0370 | 0·8830 | -1·0770 | 0·2815 |
| | Glucose levels | -0·6605 | 0·1905 | -5·4279 | -1·5079 | -3·4679 | 0·0005 |
| | CAD PRS | -0·0784 | 0·0794 | -2·9475 | 0·9725 | -0·9875 | 0·3234 |
| | LDL PRS | -0·0408 | 0·0801 | -2·4690 | 1·4510 | -0·5090 | 0·6108 |
| | SBP PRS | -0·0345 | 0·0748 | -2·4207 | 1·4993 | -0·4607 | 0·6450 |
| | DBP PRS | -0·0030 | 0·0814 | -1·9972 | 1·9228 | -0·0372 | 0·9704 |
| | SBP | 0·0571 | 0·0821 | -1·2653 | 2·6547 | 0·6947 | 0·4873 |
| | DBP | -0·0090 | 0·0727 | -2·0838 | 1·8362 | -0·1238 | 0·9015 |
| | Cholesterol levels | 0·1280 | 0·1479 | -1·0947 | 2·8253 | 0·8653 | 0·3869 |
| **TICS-m3** | AD PRS | 0·0928 | 0·0813 | -0·8189 | 3·1011 | 1·1411 | 0·2538 |
| | EA PRS | 0·1000 | 0·0666 | -0·4573 | 3·4627 | 1·5027 | 0·1329 |
| | Education years | 0·2123 | 0·0708 | 1·0360 | 4·9560 | 2·9960 | 0·0027 |
| | T2D PRS | -0·1408 | 0·0654 | -4·1123 | -0·1923 | -2·1523 | 0·0314 |
| | fg PRS | 0·0523 | 0·0772 | -1·2827 | 2·6373 | 0·6773 | 0·4982 |
| | fi PRS | -0·0966 | 0·0683 | -3·3751 | 0·5449 | -1·4151 | 0·1570 |
| | Diabetes | -0·4367 | 0·2290 | -3·8672 | 0·0528 | -1·9072 | 0·0565 |
| | Glucose levels | -0·2810 | 0·1848 | -3·4803 | 0·4397 | -1·5203 | 0·1284 |
| | CAD PRS | 0·0530 | 0·0924 | -1·3857 | 2·5343 | 0·5743 | 0·5658 |
| | LDL PRS | -0·1104 | 0·0917 | -3·1630 | 0·7570 | -1·2030 | 0·2290 |
| | SBP PRS | -0·0745 | 0·0743 | -2·9621 | 0·9579 | -1·0021 | 0·3163 |
| | DBP PRS | -0·1422 | 0·0727 | -3·9175 | 0·0025 | -1·9575 | 0·0503 |
| | SBP | 0·0073 | 0·0684 | -1·8535 | 2·0665 | 0·1065 | 0·9152 |
| | DBP | 0·0176 | 0·0798 | -1·7400 | 2·1800 | 0·2200 | 0·8259 |
| | Cholesterol levels | -0·1084 | 0·1673 | -2·6078 | 1·3122 | -0·6478 | 0·5171 |
| **GFAP** | AD PRS | 0·1046 | 0·0804 | -0·6577 | 3·2623 | 1·3023 | 0·1928 |
| | EA PRS | 0·0692 | 0·0831 | -1·1273 | 2·7927 | 0·8327 | 0·4050 |

| | | | | | | | |
|---|---|---|---|---|---|---|---|
| | Education years | 0·0815 | 0·0757 | -0·8829 | 3·0371 | 1·0771 | 0·2814 |
| | T2D PRS | -0·0469 | 0·0687 | -2·6422 | 1·2778 | -0·6822 | 0·4951 |
| | fg PRS | 0·0791 | 0·0901 | -1·0813 | 2·8387 | 0·8787 | 0·3796 |
| | fi PRS | -0·0342 | 0·0943 | -2·3228 | 1·5972 | -0·3628 | 0·7167 |
| | Diabetes | -0·3012 | 0·1665 | -3·7694 | 0·1506 | -1·8094 | 0·0704 |
| | Glucose levels | -0·3499 | 0·1507 | -4·2822 | -0·3622 | -2·3222 | 0·0202 |
| | CAD PRS | -0·0072 | 0·0890 | -2·0408 | 1·8792 | -0·0808 | 0·9356 |
| | LDL PRS | -0·0260 | 0·0790 | -2·2892 | 1·6308 | -0·3292 | 0·7420 |
| | SBP PRS | 0·0068 | 0·0715 | -1·8654 | 2·0546 | 0·0946 | 0·9246 |
| | DBP PRS | 0·0417 | 0·0702 | -1·3666 | 2·5534 | 0·5934 | 0·5529 |
| | SBP | 0·0100 | 0·0783 | -1·8325 | 2·0875 | 0·1275 | 0·8985 |
| | DBP | 0·0678 | 0·0633 | -0·8885 | 3·0315 | 1·0715 | 0·2839 |
| | Cholesterol levels | -0·1050 | 0·1375 | -2·7234 | 1·1966 | -0·7634 | 0·4452 |
| **NfL** | AD PRS | 0·0347 | 0·0586 | -1·3679 | 2·5521 | 0·5921 | 0·5538 |
| | EA PRS | 0·0425 | 0·0447 | -1·0104 | 2·9096 | 0·9496 | 0·3423 |
| | Education years | 0·0226 | 0·0552 | -1·5498 | 2·3702 | 0·4102 | 0·6816 |
| | T2D PRS | -0·0454 | 0·0593 | -2·7249 | 1·1951 | -0·7649 | 0·4443 |
| | fg PRS | 0·0093 | 0·0523 | -1·7824 | 2·1376 | 0·1776 | 0·8590 |
| | fi PRS | -0·0595 | 0·0532 | -3·0793 | 0·8407 | -1·1193 | 0·2630 |
| | Diabetes | -0·1884 | 0·1438 | -3·2700 | 0·6500 | -1·3100 | 0·1902 |
| | Glucose levels | 0·0708 | 0·1155 | -1·3472 | 2·5728 | 0·6128 | 0·5400 |
| | CAD PRS | 0·0098 | 0·0508 | -1·7673 | 2·1527 | 0·1927 | 0·8472 |
| | LDL PRS | -0·0953 | 0·0456 | -4·0508 | -0·1308 | -2·0908 | 0·0365 |
| | SBP PRS | 0·0171 | 0·0784 | -1·7417 | 2·1783 | 0·2183 | 0·8272 |
| | DBP PRS | 0·0265 | 0·1168 | -1·7334 | 2·1866 | 0·2266 | 0·8207 |
| | SBP | 0·0125 | 0·0630 | -1·7608 | 2·1592 | 0·1992 | 0·8421 |
| | DBP | -0·0490 | 0·0573 | -2·8151 | 1·1049 | -0·8551 | 0·3925 |
| | Cholesterol levels | 0·1001 | 0·2033 | -1·4678 | 2·4522 | 0·4922 | 0·6225 |

Abbreviations: SE – standard error; CI – confidence interval; GCS – Global Cognitive Score; TICS-m3 – Telephone Interview for Cognitive Status modified version 3; GFAP – glial fibrillary acidic protein; NfL – neurofilament light chain; AD – Alzheimer's disease; EA – educational attainment; T2D – type II diabetes; fg – fasting glucose; fi – fasting insulin; CAD – coronary artery disease; SBP – systolic blood pressure; DBP – diastolic blood pressure.

**Table S4. Interactions between modifiers and aging clocks significantly associated with AD-related outcomes.**

| Interactor | Outcome | Predictor | β-coefficient | SE | *p*-value |
|---|---|---|---|---|---|
| **EA PRS** | GCS | PAC | -0·0749 | 0·0675 | 0·2674 |
| | GCS | 1-HPS | -0·0205 | 0·0813 | 0·8009 |
| | GCS | Brain | -0·0784 | 0·0791 | 0·3213 |
| **Education** | GCS | PAC | -0·0287 | 0·0164 | 0·0807 |
| | GCS | 1-HPS | -0·0005 | 0·0153 | 0·9742 |
| | GCS | Brain | -0·0103 | 0·0196 | 0·5975 |
| | TICS-m3 | Brain | 0·0059 | 0·0174 | 0·7364 |
| **T2D PRS** | TICS-m3 | Brain | 0·1542 | 0·0655 | 0·0186 |
| **fg PRS** | GCS | PAC | 0·0927 | 0·0793 | 0·2429 |
| | GCS | 1-HPS | 0·1956 | 0·0952 | 0·0399 |
| | GCS | Brain | 0·1644 | 0·0879 | 0·0614 |
| **Glucose** | GCS | PAC | 0·0317 | 0·1518 | 0·8348 |
| | GCS | 1-HPS | 0·0249 | 0·1748 | 0·8866 |
| | GCS | Brain | -0·1268 | 0·1444 | 0·3800 |
| | GFAP | DunedinPACE | 0·1971 | 0·1080 | 0·0679 |
| **LDL PRS** | NFL | PAC | -0·0327 | 0·0450 | 0·4666 |
| | NFL | Conventional | -0·0242 | 0·0505 | 0·6310 |
| | NFL | Artery | -0·0357 | 0·0297 | 0·2289 |
| | NFL | Brain | -0·0423 | 0·0417 | 0·3110 |
| | NFL | Heart | 0·0234 | 0·0413 | 0·5701 |
| | NFL | Immune | 0·0231 | 0·0618 | 0·7089 |

Abbreviations: SE – standard error; PRS – polygenic risk score; EA – educational attainment; T2D – type II diabetes; fg – fasting glucose; LDL – low-density lipoprotein; GCS – Global Cognitive Score; TICS-m3 – Telephone Interview for Cognitive Status modified version 3; NfL – neurofilament light chain; PAC – Proteomic Aging Clock; HPS – Healthspan Proteomic Score.

**Table S5. Top 30 GO categories significantly enriched in each organ-specific proteomic clock.**

| Clock | Ontology | ID | Description | Fold Enrichment | Adjusted *p*-value | Gene I |
|---|---|---|---|---|---|---|
| **Adipose** | **BP** | GO:0050873 | brown fat cell differentiation | 231·39 | 0·0001 | ADIPOQ/FABP4/LEP |
| | **BP** | GO:0120162 | positive regulation of cold-induced thermogenesis | 123·71 | 0·0003 | ADIPOQ/FABP4/LEP |
| | **BP** | GO:0120161 | regulation of cold-induced thermogenesis | 82·75 | 0·0006 | ADIPOQ/FABP4/LEP |
| | **BP** | GO:0106106 | cold-induced thermogenesis | 81·67 | 0·0006 | ADIPOQ/FABP4/LEP |
| | **BP** | GO:1990845 | adaptive thermogenesis | 73·50 | 0·0006 | ADIPOQ/FABP4/LEP |
| | **BP** | GO:0001659 | temperature homeostasis | 67·54 | 0·0007 | ADIPOQ/FABP4/LEP |
| | **BP** | GO:0045444 | fat cell differentiation | 49·78 | 0·0015 | ADIPOQ/FABP4/LEP |
| | **MF** | GO:0051427 | hormone receptor binding | 245·00 | 0·0020 | FABP4/LEP |
| | **BP** | GO:0016042 | lipid catabolic process | 35·60 | 0·0032 | ADIPOQ/LEP/PLIN1 |
| | **BP** | GO:0006111 | regulation of gluconeogenesis | 151·45 | 0·0043 | ADIPOQ/LEP |
| | **BP** | GO:0046324 | regulation of D-glucose import | 138·83 | 0·0046 | ADIPOQ/LEP |
| | **BP** | GO:0032757 | positive regulation of interleukin-8 production | 128·15 | 0·0048 | ADIPOQ/LEP |
| | **BP** | GO:0006869 | lipid transport | 27·22 | 0·0048 | ADIPOQ/FABP4/LEP |
| | **BP** | GO:0046888 | negative regulation of hormone secretion | 120·72 | 0·0048 | ADIPOQ/LEP |
| | **BP** | GO:0006635 | fatty acid beta-oxidation | 112·57 | 0·0052 | ADIPOQ/LEP |
| | **BP** | GO:0010827 | regulation of D-glucose transmembrane transport | 106·79 | 0·0052 | ADIPOQ/LEP |
| | **BP** | GO:0046323 | D-glucose import | 105·44 | 0·0052 | ADIPOQ/LEP |
| | **BP** | GO:0014823 | response to activity | 101·58 | 0·0053 | ADIPOQ/LEP |
| | **BP** | GO:0006094 | gluconeogenesis | 93·60 | 0·0058 | ADIPOQ/LEP |
| | **BP** | GO:0019319 | hexose biosynthetic process | 90·54 | 0·0058 | ADIPOQ/LEP |
| | **BP** | GO:0046364 | monosaccharide biosynthetic process | 86·77 | 0·0058 | ADIPOQ/LEP |
| | **BP** | GO:0010906 | regulation of glucose metabolic process | 79·33 | 0·0058 | ADIPOQ/LEP |
| | **BP** | GO:0032637 | interleukin-8 production | 79·33 | 0·0058 | ADIPOQ/LEP |
| | **BP** | GO:0032677 | regulation of interleukin-8 production | 79·33 | 0·0058 | ADIPOQ/LEP |
| | **BP** | GO:0043255 | regulation of carbohydrate biosynthetic process | 79·33 | 0·0058 | ADIPOQ/LEP |
| | **CC** | GO:0005811 | lipid droplet | 78·58 | 0·0058 | FABP4/PLIN1 |
| | **BP** | GO:0009062 | fatty acid catabolic process | 78·58 | 0·0058 | ADIPOQ/LEP |
| | **BP** | GO:0019395 | fatty acid oxidation | 77·85 | 0·0058 | ADIPOQ/LEP |
| | **BP** | GO:0034440 | lipid oxidation | 73·07 | 0·0064 | ADIPOQ/LEP |
| | **BP** | GO:1904659 | D-glucose transmembrane | 70·59 | 0·0065 | ADIPOQ/LEP |

| | | | | | | |
|---|---|---|---|---|---|---|
| | | | transport | | | |
| **Artery** | **MF** | GO:0005201 | extracellular matrix structural constituent | 68·43 | 2·05e-08 | ACAN/BGN/ELN/LTBP2/MFGE8/THBS2 |
| | **MF** | GO:0005539 | glycosaminoglycan binding | 46·18 | 1·10e-07 | ACAN/BGN/LTBP2/SUSD5/THBS2/TNFRSF11B |
| | **CC** | GO:0062023 | collagen-containing extracellular matrix | 26·42 | 2·07e-06 | ACAN/BGN/ELN/LTBP2/MFGE8/THBS2 |
| | **MF** | GO:0050840 | extracellular matrix binding | 87·34 | 0·0002 | BGN/ELN/LTBP2 |
| | **MF** | GO:0030021 | extracellular matrix structural constituent conferring compression resistance | 172·11 | 0·0023 | ACAN/BGN |
| | **MF** | GO:0005540 | hyaluronic acid binding | 157·77 | 0·0023 | ACAN/SUSD5 |
| | **MF** | GO:1901681 | sulfur compound binding | 20·50 | 0·0099 | LTBP2/THBS2/TNFRSF11B |
| | **CC** | GO:0043202 | lysosomal lumen | 38·64 | 0·0287 | ACAN/BGN |
| | **CC** | GO:0005796 | Golgi lumen | 35·72 | 0·0298 | ACAN/BGN |
| | **MF** | GO:0019955 | cytokine binding | 26·48 | 0·0485 | BGN/CRLF1 |
| **Brain** | **BP** | GO:0007626 | locomotory behavior | 13·04 | 4·86e-06 | CEND1/CNP/CNTN1/CNTN2/CRH/GPR37/PENK/SEZ6/SNAP25/TNR |
| | **BP** | GO:0060074 | synapse maturation | 41·65 | 4·86e-06 | BCAN/C1QL2/IGSF21/NEFL/SEZ6/SEZ6L |
| | **BP** | GO:0007409 | axonogenesis | 7·86 | 4·86e-06 | APLP1/CNP/CNTN1/CNTN2/LRTM2/MAP2/NEFL/NPTX1/PTPRZ1/RTN4R/SLITRK1/SNAP25/TNR |
| | **BP** | GO:0050807 | regulation of synapse organization | 9·79 | 5·94e-06 | C1QL2/DNM3/GPR158/GRIN2B/LRTM2/MDGA1/NEFL/OXT/RTN4R/SLITRK1/SNAP25 |
| | **BP** | GO:0050803 | regulation of synapse structure or activity | 9·60 | 5·94e-06 | C1QL2/DNM3/GPR158/GRIN2B/LRTM2/MDGA1/NEFL/OXT/RTN4R/SLITRK1/SNAP25 |
| | **CC** | GO:0072534 | perineuronal net | 111·07 | 8·72e-06 | BCAN/NCAN/PTPRZ1/TNR |
| | **CC** | GO:0098966 | perisynaptic extracellular matrix | 100·97 | 1·17e-05 | BCAN/NCAN/PTPRZ1/TNR |
| | **BP** | GO:0010975 | regulation of neuron projection development | 7·37 | 1·23e-05 | CNTN1/CNTN2/DNM3/GFAP/MAP2/NEFL/RTN4R/SEZ6/SLITRK1/SNAP25/SPOCK1/TNR |
| | **CC** | GO:0097060 | synaptic membrane | 7·36 | 1·23e-05 | CNTN1/CNTN2/CNTNAP2/CNTNAP4/DNM1/DNM3/GPR158/GRIN2B/IGSF21/SLITRK1/SNAP25/SYT1 |
| | **CC** | GO:0099535 | synapse-associated extracellular matrix | 92·56 | 1·23e-05 | BCAN/NCAN/PTPRZ1/TNR |
| | **BP** | GO:0031102 | neuron projection regeneration | 27·31 | 1·25e-05 | APOD/GFAP/NEFL/OMG/RTN4R/TNR |
| | **BP** | GO:0050806 | positive regulation of synaptic transmission | 12·62 | 3·14e-05 | CRH/GFAP/GPR158/GRIN2B/OXT/SNAP25/SYT1/TNR |
| | **CC** | GO:0044304 | main axon | 22·82 | 3·14e-05 | CNTN2/CNTNAP2/CRH/MAP2/SCN2B/SPOCK1 |
| | **CC** | GO:0042734 | presynaptic membrane | 11·94 | 4·11e-05 | CNTN1/CNTNAP2/CNTNAP4/DNM1/GPR158/IGSF21/SNAP25/SYT1 |
| | **BP** | GO:0021700 | developmental maturation | 7·93 | 5·33e-05 | ATP6V1G2/BCAN/C1QL2/CEND1/CNTN2/CNTNAP2/IGSF21/NEFL/SEZ6/SEZ6L |

| | | | | | | |
|---|---|---|---|---|---|---|
| | **BP** | GO:0007416 | synapse assembly | 9·29 | 5·33e-05 | C1QL2/DNM3/LRTM2/MDGA1/NPTX1/OXT/RTN4R/SLITRK1/SNAP25 |
| | **BP** | GO:0051960 | regulation of nervous system development | 6·60 | 7·61e-05 | BCAN/GFAP/LRTM2/MAP2/NCAN/NEFL/OXT/PTPRZ1/RTN4R/SLITRK1/TNR |
| | **CC** | GO:0098984 | neuron to neuron synapse | 6·74 | 0·0002 | C1QL2/DNM3/GPR158/GRIN2B/IGSF21/PENK/PPP3R1/SLITRK1/SPOCK1/SYT1 |
| | **BP** | GO:0030900 | forebrain development | 6·67 | 0·0002 | APLP1/BCAN/CNP/CNTN2/CNTNAP2/CRH/MDGA1/NEFL/RTN4R/TNR |
| | **BP** | GO:0050890 | cognition | 7·60 | 0·0002 | CNTN2/CNTNAP2/CRH/GPR158/GRIN2B/OXT/PTPRZ1/SNAP25/TNR |
| | **BP** | GO:0042063 | gliogenesis | 7·04 | 0·0004 | BCAN/CNP/CNTN1/CNTN2/GFAP/NCAN/PENK/PPP3R1/PTPRZ1 |
| | **BP** | GO:0031345 | negative regulation of cell projection organization | 10·34 | 0·0004 | DNM3/GFAP/GRIN2B/MAP2/RTN4R/SPOCK1/TNR |
| | **BP** | GO:0010001 | glial cell differentiation | 8·32 | 0·0004 | BCAN/CNP/CNTN1/CNTN2/GFAP/NCAN/PPP3R1/PTPRZ1 |
| | **BP** | GO:1901888 | regulation of cell junction assembly | 8·29 | 0·0004 | APOD/CNTNAP2/LRTM2/MDGA1/OXT/RTN4R/SLITRK1/SNAP25 |
| | **BP** | GO:0007611 | learning or memory | 7·88 | 0·0005 | CNTN2/CNTNAP2/CRH/GRIN2B/OXT/PTPRZ1/SNAP25/TNR |
| | **BP** | GO:0010977 | negative regulation of neuron projection development | 12·62 | 0·0005 | DNM3/GFAP/MAP2/RTN4R/SPOCK1/TNR |
| | **BP** | GO:0050767 | regulation of neurogenesis | 6·52 | 0·0006 | BCAN/GFAP/MAP2/NCAN/NEFL/PTPRZ1/RTN4R/SLITRK1/TNR |
| | **BP** | GO:0051962 | positive regulation of nervous system development | 7·48 | 0·0007 | BCAN/GFAP/LRTM2/NCAN/NEFL/OXT/PTPRZ1/SLITRK1 |
| | **BP** | GO:0048167 | regulation of synaptic plasticity | 8·83 | 0·0008 | CNTN2/CRH/GFAP/GRIN2B/PENK/SNAP25/TNR |
| | **BP** | GO:0050770 | regulation of axonogenesis | 11·41 | 0·0008 | CNTN2/MAP2/NEFL/RTN4R/SLITRK1/TNR |
| **Heart** | **CC** | GO:0030017 | sarcomere | 53·60 | 6·25e-05 | BMP10/ITGB1BP2/MYL4/TNNI3 |
| | **CC** | GO:0030016 | myofibril | 48·97 | 6·25e-05 | BMP10/ITGB1BP2/MYL4/TNNI3 |
| | **CC** | GO:0043292 | contractile muscle fiber | 47·04 | 6·25e-05 | BMP10/ITGB1BP2/MYL4/TNNI3 |
| | **BP** | GO:0060048 | cardiac muscle contraction | 60·71 | 0·0008 | BMP10/MYL4/TNNI3 |
| | **BP** | GO:0006941 | striated muscle contraction | 46·01 | 0·0014 | BMP10/MYL4/TNNI3 |
| | **BP** | GO:0008016 | regulation of heart contraction | 41·90 | 0·0015 | BMP10/MYL4/TNNI3 |
| | **BP** | GO:0048738 | cardiac muscle tissue development | 35·42 | 0·0018 | BMP10/NPPB/TNNI3 |
| | **BP** | GO:0060047 | heart contraction | 35·28 | 0·0018 | BMP10/MYL4/TNNI3 |
| | **BP** | GO:0003015 | heart process | 34·06 | 0·0018 | BMP10/MYL4/TNNI3 |
| | **BP** | GO:1903522 | regulation of blood circulation | 33·43 | 0·0018 | BMP10/MYL4/TNNI3 |
| | **BP** | GO:0055010 | ventricular cardiac muscle tissue morphogenesis | 126·60 | 0·0024 | BMP10/TNNI3 |
| | **BP** | GO:0006936 | muscle contraction | 24·93 | 0·0030 | BMP10/MYL4/TNNI3 |
| | **BP** | GO:0055008 | cardiac muscle tissue morphogenesis | 99·17 | 0·0030 | BMP10/TNNI3 |

| | | | | | | |
|---|---|---|---|---|---|---|
| | **BP** | GO:0007517 | muscle organ development | 24·19 | 0·0030 | BMP10/ITGB1BP2/TNNI3 |
| | **BP** | GO:0003229 | ventricular cardiac muscle tissue development | 94·44 | 0·0030 | BMP10/TNNI3 |
| | **BP** | GO:0043462 | regulation of ATP-dependent activity | 94·44 | 0·0030 | MYL4/TNNI3 |
| | **BP** | GO:0003208 | cardiac ventricle morphogenesis | 83·80 | 0·0036 | BMP10/TNNI3 |
| | **BP** | GO:0060415 | muscle tissue morphogenesis | 81·51 | 0·0036 | BMP10/TNNI3 |
| | **BP** | GO:0014706 | striated muscle tissue development | 21·05 | 0·0038 | BMP10/NPPB/TNNI3 |
| | **BP** | GO:0055117 | regulation of cardiac muscle contraction | 75·32 | 0·0038 | BMP10/TNNI3 |
| | **BP** | GO:0048644 | muscle organ morphogenesis | 73·46 | 0·0038 | BMP10/TNNI3 |
| | **BP** | GO:0060537 | muscle tissue development | 19·97 | 0·0038 | BMP10/NPPB/TNNI3 |
| | **BP** | GO:0003012 | muscle system process | 19·19 | 0·0041 | BMP10/MYL4/TNNI3 |
| | **BP** | GO:0003073 | regulation of systemic arterial blood pressure | 60·71 | 0·0049 | NPPB/TNNI3 |
| | **BP** | GO:0006942 | regulation of striated muscle contraction | 58·91 | 0·0050 | BMP10/TNNI3 |
| | **BP** | GO:0003231 | cardiac ventricle development | 46·48 | 0·0074 | BMP10/TNNI3 |
| | **BP** | GO:0003206 | cardiac chamber morphogenesis | 46·12 | 0·0074 | BMP10/TNNI3 |
| | **MF** | GO:0005179 | hormone activity | 45·08 | 0·0074 | BMP10/NPPB |
| | **CC** | GO:0030018 | Z disc | 44·74 | 0·0074 | BMP10/ITGB1BP2 |
| | **CC** | GO:0031674 | I band | 40·20 | 0·0089 | BMP10/ITGB1BP2 |
| **Immune** | **CC** | GO:0070820 | tertiary granule | 23·90 | 8·23e-15 | CD177/CD300A/CEACAM8/CFP/CLEC12A/CLEC4D/CXCL1/FOLR3/ITGAM/LILRB2/MCEMP1/MMP8/MMP9/PGLYRP1/PRG3/SIGLEC5 |
| | **CC** | GO:0042581 | specific granule | 22·97 | 8·96e-14 | BST1/CD177/CEACAM8/CFP/CLEC12A/CLEC4D/CXCL1/FOLR3/ITGAM/MCEMP1/MMP8/PGLYRP1/PRG3/RETN/TNFRSF1B |
| | **BP** | GO:0001906 | cell killing | 15·78 | 2·67e-13 | AZU1/CD5L/CORO1A/CXCL1/CXCL13/CXCL6/FCGR2A/FCGR3B/GZMA/GZMB/GZMH/IL18RAP/ITGAM/PF4/PGLYRP1/PTPRC/S100A12 |
| | **BP** | GO:0002274 | myeloid leukocyte activation | 15·87 | 1·44e-12 | ADGRE2/AIF1/AZU1/CCL5/CD177/CD300A/CLEC4D/CST7/CXCL6/IL18RAP/ITGAM/MMP8/PRG3/PTPRC/S100A12/TLR1 |
| | **CC** | GO:0030667 | secretory granule membrane | 13·02 | 3·89e-12 | AZU1/BST1/CD177/CD300A/CEACAM8/CLEC12A/CLEC4D/FCGR2A/FCGR3B/ITGAM/LILRB2/MCEMP1/PTPRC/SELL/SIGLEC5/SIRPB1/TNFRSF1B |
| | **CC** | GO:0060205 | cytoplasmic vesicle lumen | 12·66 | 4·60e-12 | AZU1/BIN2/CFP/CXCL1/FASLG/FCN1/FOLR3/GZMB/MMP8/MPO/PF4/PGLYRP1/PRG3/PRTN3/RETN/RNASE3/S100A12 |
| | **CC** | GO:0031983 | vesicle lumen | 12·62 | 4·60e-12 | AZU1/BIN2/CFP/CXCL1/FASLG/FCN1/FOLR3/GZMB/MMP8/MPO/PF4/PGLYRP1/PRG3/PRTN3/RETN/RNASE3/S100A12 |

| | | | | | | |
|---|---|---|---|---|---|---|
| | **BP** | GO:0050900 | leukocyte migration | 10·92 | 7·69e-12 | ADGRE2/AIF1/AZU1/BST1/CCL5/CD177/CD300A/CORO1A/CSF1R/CSF3R/CXCL1/CXCL13/CXCL6/PF4/PRTN3/S100A12/SELL/SELPLG |
| | **BP** | GO:0006909 | phagocytosis | 15·38 | 8·01e-12 | AIF1/AZU1/BIN2/CD300A/CFP/CORO1A/FCGR2A/FCN1/ICAM3/IL2RG/ITGAM/NCF2/PRTN3/PTPRC/SIRPB1 |
| | **BP** | GO:0097529 | myeloid leukocyte migration | 15·12 | 9·19e-12 | ADGRE2/AIF1/AZU1/BST1/CCL5/CD177/CD300A/CSF1R/CSF3R/CXCL1/CXCL13/CXCL6/PF4/PRTN3/S100A12 |
| | **BP** | GO:0097530 | granulocyte migration | 20·42 | 9·57e-12 | ADGRE2/BST1/CCL5/CD177/CD300A/CSF1R/CSF3R/CXCL1/CXCL13/CXCL6/PF4/PRTN3/S100A12 |
| | **BP** | GO:0006959 | humoral immune response | 11·85 | 2·63e-10 | AZU1/BST1/CCL5/CD5L/CFP/CXCL1/CXCL13/CXCL6/FCN1/PF4/PGLYRP1/PRTN3/PTPRC/RNASE3/S100A12 |
| | **CC** | GO:0034774 | secretory granule lumen | 11·34 | 4·57e-10 | AZU1/BIN2/CFP/CXCL1/FCN1/FOLR3/MMP8/MPO/PF4/PGLYRP1/PRG3/PRTN3/RETN/RNASE3/S100A12 |
| | **BP** | GO:0002697 | regulation of immune effector process | 9·56 | 9·97e-10 | ADGRE2/CD177/CD300A/CD5L/CD7/CFP/CLC/CXCL6/FCN1/IL18RAP/ITGAM/MZB1/PGLYRP1/PRG2/PTPRC/TNFRSF1B |
| | **BP** | GO:0002443 | leukocyte mediated immunity | 8·59 | 1·05e-09 | ADGRE2/AZU1/CD177/CD300A/CD7/CD8A/CLC/CORO1A/CXCL6/FCGR2A/FCGR3B/GZMB/IL18RAP/ITGAM/PTPRC/SLA2/TNFRSF1B |
| | **CC** | GO:0070821 | tertiary granule membrane | 30·21 | 1·53e-09 | CD177/CD300A/CEACAM8/CLEC12A/CLEC4D/ITGAM/LILRB2/MCEMP1/SIGLEC5 |
| | **BP** | GO:0030595 | leukocyte chemotaxis | 13·27 | 1·53e-09 | ADGRE2/AIF1/AZU1/BST1/CCL5/CORO1A/CSF1R/CSF3R/CXCL1/CXCL13/CXCL6/PF4/S100A12 |
| | **BP** | GO:0060326 | cell chemotaxis | 10·45 | 5·44e-09 | ADGRE2/AIF1/AZU1/BIN2/BST1/CCL5/CORO1A/CSF1R/CSF3R/CXCL1/CXCL13/CXCL6/PF4/S100A12 |
| | **BP** | GO:0006935 | chemotaxis | 8·31 | 5·98e-09 | ADGRE2/AIF1/AZU1/BIN2/BST1/CCL5/CORO1A/CSF1R/CSF3R/CXCL1/CXCL13/CXCL6/LRTM1/PF4/RNASE3/S100A12 |
| | **BP** | GO:0042330 | taxis | 8·27 | 6·04e-09 | ADGRE2/AIF1/AZU1/BIN2/BST1/CCL5/CORO1A/CSF1R/CSF3R/CXCL1/CXCL13/CXCL6/LRTM1/PF4/RNASE3/S100A12 |
| | **BP** | GO:0031640 | killing of cells of another organism | 19·76 | 6·09e-09 | AZU1/CXCL1/CXCL13/CXCL6/GZMA/GZMB/GZMH/PF4/PGLYRP1/S100A12 |
| | **BP** | GO:0141061 | disruption of cell in another organism | 19·76 | 6·09e-09 | AZU1/CXCL1/CXCL13/CXCL6/GZMA/GZMB/GZMH/PF4/PGLYRP1/S100A12 |
| | **CC** | GO:0009897 | external side of plasma membrane | 9·03 | 6·49e-09 | ADGRE1/CD8A/CEACAM21/CLEC4D/CSF2RA/CSF3R/FASLG/FCGR2A/FCGR3B/FCN1/FOLR3/IL2RG/ITGAM/PTPRC/SELL |
| | **BP** | GO:0141060 | disruption of anatomical structure in another organism | 19·29 | 7·08e-09 | AZU1/CXCL1/CXCL13/CXCL6/GZMA/GZMB/GZMH/PF4/PGLYRP1/S100A12 |
| | **BP** | GO:0071621 | granulocyte chemotaxis | 18·99 | 7·94e-09 | ADGRE2/BST1/CCL5/CSF1R/CSF3R/CXCL1/CXCL13/CXCL6/PF4/S100A12 |
| | **BP** | GO:0050764 | regulation of phagocytosis | 21·62 | 2·12e-08 | AZU1/CD300A/CFP/FCGR2A/FCN1/IL2RG/PRTN3/PTPRC/SIRPB1 |
| | **BP** | GO:0042119 | neutrophil activation | 38·98 | 2·96e-08 | CCL5/CD177/CD300A/CXCL6/IL18RAP/ITGAM/PRG3 |

| | | | | | | |
|---|---|---|---|---|---|---|
| | **BP** | GO:1990266 | neutrophil migration | 20·42 | 3·30e-08 | BST1/CD177/CSF3R/CXCL1/CXCL13/CXCL6/PF4/PRTN3/S100A12 |
| | **MF** | GO:0140375 | immune receptor activity | 15·91 | 3·93e-08 | CSF2RA/CSF3R/FCGR2A/FCGR3B/IL18RAP/IL1R2/IL2RG/KLRF1/LILRA5/LILRB2 |
| | **BP** | GO:0070663 | regulation of leukocyte proliferation | 10·89 | 5·38e-08 | AIF1/BST1/CCL5/CD300A/CLC/CORO1A/CSF1R/CSF2RA/LILRB2/MZB1/PTPRC/TNFRSF1B |
| **Intestine** | **MF** | GO:0005179 | hormone activity | 27·20 | 0·0004 | CCL25/GUCA2A/INSL5/MLN/PYY |
| | **CC** | GO:0005902 | microvillus | 28·72 | 0·0017 | CDHR2/CDHR5/CEACAM20/FABP2 |
| | **CC** | GO:0031528 | microvillus membrane | 65·28 | 0·0017 | CDHR2/CDHR5/CEACAM20 |
| | **BP** | GO:0022600 | digestive system process | 25·65 | 0·00173 | APOA4/FABP2/MUC13/MUC2 |
| | **MF** | GO:0005504 | fatty acid binding | 45·84 | 0·0027 | FABP2/FABP6/RBP2 |
| | **CC** | GO:0045177 | apical part of cell | 9·09 | 0·0027 | CDHR2/CDHR5/CEACAM20/FABP2/MUC13/S100G |
| | **BP** | GO:0007586 | digestion | 19·95 | 0·0027 | APOA4/FABP2/MUC13/MUC2 |
| | **BP** | GO:0032530 | regulation of microvillus organization | 110·48 | 0·0073 | CDHR2/CDHR5 |
| | **BP** | GO:0032536 | regulation of cell projection size | 102·59 | 0·0073 | CDHR2/CDHR5 |
| | **MF** | GO:0033293 | monocarboxylic acid binding | 27·27 | 0·0073 | FABP2/FABP6/RBP2 |
| | **CC** | GO:0016324 | apical plasma membrane | 8·80 | 0·0085 | CDHR2/CDHR5/CEACAM20/MUC13/S100G |
| | **CC** | GO:0098858 | actin-based cell projection | 12·28 | 0·0101 | CDHR2/CDHR5/CEACAM20/FABP2 |
| | **CC** | GO:0005903 | brush border | 19·76 | 0·0130 | CDHR2/CDHR5/TMPRSS15 |
| | **BP** | GO:0030277 | maintenance of gastrointestinal epithelium | 59·84 | 0·0130 | MUC13/MUC2 |
| | **BP** | GO:0032528 | microvillus organization | 59·84 | 0·0130 | CDHR2/CDHR5 |
| | **BP** | GO:0098856 | intestinal lipid absorption | 59·84 | 0·0130 | APOA4/FABP2 |
| | **BP** | GO:0015908 | fatty acid transport | 17·95 | 0·0147 | FABP2/FABP6/RBP2 |
| | **MF** | GO:0001664 | G protein-coupled receptor binding | 9·61 | 0·0170 | CCL25/INSL5/MLN/PYY |
| | **BP** | GO:0010669 | epithelial structure maintenance | 37·79 | 0·0275 | MUC13/MUC2 |
| | **BP** | GO:0007156 | homophilic cell adhesion via plasma membrane adhesion molecules | 12·82 | 0·0327 | CDH17/CDHR2/CDHR5 |
| | **BP** | GO:0050892 | intestinal absorption | 32·64 | 0·0327 | APOA4/FABP2 |
| | **CC** | GO:0098862 | cluster of actin-based cell projections | 12·45 | 0·0327 | CDHR2/CDHR5/TMPRSS15 |
| | **BP** | GO:0015718 | monocarboxylic acid transport | 12·04 | 0·0345 | FABP2/FABP6/RBP2 |
| | **MF** | GO:0031406 | carboxylic acid binding | 11·52 | 0·0361 | FABP2/FABP6/RBP2 |
| | **BP** | GO:0044331 | cell-cell adhesion mediated by cadherin | 28·72 | 0·0361 | CDH17/CDHR2 |
| | **MF** | GO:0043177 | organic acid binding | 10·83 | 0·0412 | FABP2/FABP6/RBP2 |

| | | | | | | |
|---|---|---|---|---|---|---|
| | **CC** | GO:0031526 | brush border membrane | 23·94 | 0·0479 | CDHR2/CDHR5 |
| **Kidney** | **BP** | GO:0003014 | renal system process | 78·88 | 0·0190 | REN/UMOD |
| | **BP** | GO:0008217 | regulation of blood pressure | 54·80 | 0·0190 | REN/UMOD |
| | **BP** | GO:0009755 | hormone-mediated signaling pathway | 46·69 | 0·0190 | CYP24A1/REN |
| | **BP** | GO:0001822 | kidney development | 32·34 | 0·0190 | REN/UMOD |
| | **BP** | GO:0072001 | renal system development | 31·37 | 0·0190 | REN/UMOD |
| | **BP** | GO:0032496 | response to lipopolysaccharide | 29·41 | 0·0190 | REN/UMOD |
| | **BP** | GO:0002237 | response to molecule of bacterial origin | 27·77 | 0·0190 | REN/UMOD |
| | **BP** | GO:0002002 | regulation of angiotensin levels in blood | 520·63 | 0·0190 | REN |
| | **BP** | GO:0030002 | intracellular monoatomic anion homeostasis | 520·63 | 0·0190 | UMOD |
| | **BP** | GO:0030643 | intracellular phosphate ion homeostasis | 520·63 | 0·0190 | UMOD |
| | **BP** | GO:0030644 | intracellular chloride ion homeostasis | 520·63 | 0·0190 | UMOD |
| | **BP** | GO:0140367 | antibacterial innate immune response | 520·63 | 0·0190 | UMOD |
| | **BP** | GO:1903044 | protein localization to membrane raft | 520·63 | 0·0190 | UMOD |
| | **BP** | GO:0015747 | urate transport | 473·30 | 0·0190 | UMOD |
| | **MF** | GO:0019864 | IgG binding | 473·30 | 0·0190 | UMOD |
| | **BP** | GO:0042756 | drinking behavior | 473·30 | 0·0190 | REN |
| | **BP** | GO:0071918 | urea transmembrane transport | 473·30 | 0·0190 | UMOD |
| | **BP** | GO:0015840 | urea transport | 433·85 | 0·0190 | UMOD |
| | **BP** | GO:0042363 | fat-soluble vitamin catabolic process | 433·85 | 0·0190 | CYP24A1 |
| | **BP** | GO:0072017 | distal tubule development | 433·85 | 0·0190 | UMOD |
| | **BP** | GO:0072070 | loop of Henle development | 433·85 | 0·0190 | UMOD |
| | **BP** | GO:0009415 | response to water | 400·48 | 0·0190 | UMOD |
| | **BP** | GO:0070294 | renal sodium ion absorption | 400·48 | 0·0190 | UMOD |
| | **BP** | GO:0070561 | vitamin D receptor signaling pathway | 400·48 | 0·0190 | CYP24A1 |
| | **BP** | GO:0009111 | vitamin catabolic process | 371·88 | 0·0190 | CYP24A1 |
| | **BP** | GO:0009410 | response to xenobiotic stimulus | 23·09 | 0·0190 | REN/UMOD |
| | **BP** | GO:0072044 | collecting duct development | 347·08 | 0·0190 | UMOD |
| | **CC** | GO:0045177 | apical part of cell | 21·97 | 0·0190 | REN/UMOD |
| | **BP** | GO:0003096 | renal sodium ion transport | 325·39 | 0·0190 | UMOD |
| | **MF** | GO:0005159 | insulin-like growth factor receptor binding | 325·39 | 0·0190 | REN |

| | | | | | | |
|---|---|---|---|---|---|---|
| **Liver** | **CC** | GO:0072562 | blood microparticle | 39·02 | 8·29e-20 | A1BG/AFM/AGT/AHSG/AMBP/APCS/APOA2/C9/CFB/FGA/GC/HRG/ORM1/PLG/PON1/PZP/SERPINF2 |
| | **BP** | GO:0007596 | blood coagulation | 24·01 | 1·61e-16 | C4BPB/CPB2/F11/F12/F13B/F7/F9/FGA/FGL1/HGFAC/HRG/KLKB1/PLG/PROC/SERPINA1/SERPIND1/SERPINF2 |
| | **BP** | GO:0007599 | hemostasis | 23·32 | 1·61e-16 | C4BPB/CPB2/F11/F12/F13B/F7/F9/FGA/FGL1/HGFAC/HRG/KLKB1/PLG/PROC/SERPINA1/SERPIND1/SERPINF2 |
| | **BP** | GO:0050817 | coagulation | 23·32 | 1·61e-16 | C4BPB/CPB2/F11/F12/F13B/F7/F9/FGA/FGL1/HGFAC/HRG/KLKB1/PLG/PROC/SERPINA1/SERPIND1/SERPINF2 |
| | **CC** | GO:0062023 | collagen-containing extracellular matrix | 14·61 | 5·17e-15 | A1BG/AGT/AHSG/AMBP/ANG/ANGPTL3/APCS/F12/F7/F9/FGA/FGL1/HRG/MST1/ORM1/PLG/PZP/SERPINA1/SERPINF2 |
| | **BP** | GO:0042060 | wound healing | 13·40 | 1·58e-13 | APCS/C4BPB/CPB2/F11/F12/F13B/F7/F9/FGA/FGL1/HGFAC/HRG/KLKB1/PLG/PROC/SERPINA1/SERPIND1/SERPINF2 |
| | **BP** | GO:0050878 | regulation of body fluid levels | 14·75 | 2·04e-13 | C4BPB/CPB2/F11/F12/F13B/F7/F9/FGA/FGL1/HGFAC/HRG/KLKB1/PLG/PROC/SERPINA1/SERPIND1/SERPINF2 |
| | **MF** | GO:0004866 | endopeptidase inhibitor activity | 25·58 | 5·30e-13 | AGT/AHSG/AMBP/FETUB/HRG/ITIH3/LPA/PZP/SERPINA1/SERPINA11/SERPINA6/SERPIND1/SERPINF2 |
| | **MF** | GO:0030414 | peptidase inhibitor activity | 24·42 | 8·67e-13 | AGT/AHSG/AMBP/FETUB/HRG/ITIH3/LPA/PZP/SERPINA1/SERPINA11/SERPINA6/SERPIND1/SERPINF2 |
| | **BP** | GO:0030193 | regulation of blood coagulation | 47·91 | 1·28e-12 | CPB2/F11/F12/F7/FGA/HRG/KLKB1/PLG/PROC/SERPINF2 |
| | **BP** | GO:0042730 | fibrinolysis | 97·94 | 1·28e-12 | CPB2/F11/F12/FGA/HRG/KLKB1/PLG/SERPINF2 |
| | **BP** | GO:1900046 | regulation of hemostasis | 46·56 | 1·52e-12 | CPB2/F11/F12/F7/FGA/HRG/KLKB1/PLG/PROC/SERPINF2 |
| | **MF** | GO:0061135 | endopeptidase regulator activity | 22·74 | 1·52e-12 | AGT/AHSG/AMBP/FETUB/HRG/ITIH3/LPA/PZP/SERPINA1/SERPINA11/SERPINA6/SERPIND1/SERPINF2 |
| | **BP** | GO:0050818 | regulation of coagulation | 43·49 | 2·66e-12 | CPB2/F11/F12/F7/FGA/HRG/KLKB1/PLG/PROC/SERPINF2 |
| | **BP** | GO:0030195 | negative regulation of blood coagulation | 59·50 | 2·66e-12 | CPB2/F11/F12/FGA/HRG/KLKB1/PLG/PROC/SERPINF2 |
| | **BP** | GO:1900047 | negative regulation of hemostasis | 58·33 | 2·87e-12 | CPB2/F11/F12/FGA/HRG/KLKB1/PLG/PROC/SERPINF2 |
| | **BP** | GO:0061045 | negative regulation of wound healing | 42·38 | 2·87e-12 | APCS/CPB2/F11/F12/FGA/HRG/KLKB1/PLG/PROC/SERPINF2 |
| | **BP** | GO:0051917 | regulation of fibrinolysis | 128·55 | 4·25e-12 | CPB2/F11/F12/HRG/KLKB1/PLG/SERPINF2 |
| | **BP** | GO:0050819 | negative regulation of coagulation | 55·09 | 4·42e-12 | CPB2/F11/F12/FGA/HRG/KLKB1/PLG/PROC/SERPINF2 |
| | **MF** | GO:0004857 | enzyme inhibitor activity | 12·90 | 4·50e-12 | AGT/AHSG/AMBP/ANGPTL3/APOA2/APOC1/FETUB/HRG/ITIH3/LPA/PZP/SERPINA1/SERPINA11/SERPINA6/SERPIND1/SERPINF2 |
| | **MF** | GO:0061134 | peptidase regulator activity | 18·29 | 1·57e-11 | AGT/AHSG/AMBP/FETUB/HRG/ITIH3/LPA/PZP/SERPINA1/SERPINA11/SERPINA6/SERPIND1/SERPINF2 |
| | **BP** | GO:0061041 | regulation of wound healing | 25·97 | 2·65e-11 | APCS/CPB2/F11/F12/F7/FGA/HRG/KLKB1/PLG/PROC/SERPINF2 |

| | | | | | | |
|---|---|---|---|---|---|---|
| | **BP** | GO:1903035 | negative regulation of response to wounding | 32·73 | 3·08e-11 | APCS/CPB2/F11/F12/FGA/HRG/KLKB1/PLG/PROC/SERPINF2 |
| | **MF** | GO:0004867 | serine-type endopeptidase inhibitor activity | 32·41 | 3·21e-11 | AGT/AMBP/HRG/ITIH3/PZP/SERPINA1/SERPINA11/SERPINA6/SERPIND1/SERPINF2 |
| | **BP** | GO:0051346 | negative regulation of hydrolase activity | 17·05 | 3·21e-11 | AMBP/ANGPTL3/APCS/APOA2/APOC1/FETUB/ITIH3/PZP/SERPINA1/SERPINA11/SERPINA6/SERPIND1/SERPINF2 |
| | **BP** | GO:0002526 | acute inflammatory response | 30·05 | 6·50e-11 | AHSG/APCS/EPO/F12/KLKB1/LBP/MBL2/ORM1/SERPINA1/SERPINF2 |
| | **BP** | GO:0006953 | acute-phase response | 55·09 | 8·44e-11 | AHSG/APCS/EPO/LBP/MBL2/ORM1/SERPINA1/SERPINF2 |
| | **BP** | GO:1903034 | regulation of response to wounding | 20·09 | 3·51e-10 | APCS/CPB2/F11/F12/F7/FGA/HRG/KLKB1/PLG/PROC/SERPINF2 |
| | **BP** | GO:0031638 | zymogen activation | 45·59 | 3·90e-10 | CPB2/F11/F12/F9/FGA/HGFAC/KLKB1/SERPINF2 |
| | **CC** | GO:0031093 | platelet alpha granule lumen | 39·47 | 1·26e-09 | A1BG/AHSG/FGA/HRG/ORM1/PLG/SERPINA1/SERPINF2 |
| **Pancreas** | **BP** | GO:0007586 | digestion | 38·94 | 1·51e-07 | AMY2A/CLPS/CTRB1/PNLIP/PNLIPRP2/PRSS2/SPINK1 |
| | **MF** | GO:0004252 | serine-type endopeptidase activity | 31·50 | 3·34e-07 | CELA2A/CELA3A/CTRB1/CTRC/CTRL/KLK1/PRSS2 |
| | **MF** | GO:0008236 | serine-type peptidase activity | 28·75 | 3·63e-07 | CELA2A/CELA3A/CTRB1/CTRC/CTRL/KLK1/PRSS2 |
| | **MF** | GO:0017171 | serine hydrolase activity | 28·17 | 3·63e-07 | CELA2A/CELA3A/CTRB1/CTRC/CTRL/KLK1/PRSS2 |
| | **MF** | GO:0004175 | endopeptidase activity | 13·88 | 3·69e-05 | CELA2A/CELA3A/CTRB1/CTRC/CTRL/KLK1/PRSS2 |
| | **BP** | GO:0043434 | response to peptide hormone | 12·57 | 5·70e-05 | CELA2A/GCG/PLA2G1B/REG1A/REG1B/REG3G/SPINK1 |
| | **CC** | GO:0042589 | zymogen granule membrane | 141·35 | 5·70e-05 | CUZD1/GP2/PNLIPRP2 |
| | **MF** | GO:0070492 | oligosaccharide binding | 141·35 | 5·70e-05 | REG1A/REG1B/REG3G |
| | **CC** | GO:0042588 | zymogen granule | 126·47 | 6·49e-05 | CUZD1/GP2/PNLIPRP2 |
| | **MF** | GO:0042834 | peptidoglycan binding | 126·47 | 6·49e-05 | REG1A/REG1B/REG3G |
| | **BP** | GO:0019730 | antimicrobial humoral response | 21·65 | 0·0001 | PLA2G1B/PRSS2/REG1A/REG1B/REG3G |
| | **MF** | GO:0004806 | triacylglycerol lipase activity | 100·12 | 0·0001 | PNLIP/PNLIPRP1/PNLIPRP2 |
| | **MF** | GO:0016298 | lipase activity | 24·84 | 0·0006 | PLA2G1B/PNLIP/PNLIPRP1/PNLIPRP2 |
| | **BP** | GO:0061844 | antimicrobial humoral immune response mediated by antimicrobial peptide | 23·56 | 0·0007 | PLA2G1B/REG1A/REG1B/REG3G |
| | **BP** | GO:0006959 | humoral immune response | 12·92 | 0·0009 | PLA2G1B/PRSS2/REG1A/REG1B/REG3G |
| | **BP** | GO:0009306 | protein secretion | 10·79 | 0·0020 | CELA2A/GCG/PLA2G1B/PPY/SEL1L |
| | **BP** | GO:0035592 | establishment of protein localization to extracellular region | 10·71 | 0·0020 | CELA2A/GCG/PLA2G1B/PPY/SEL1L |
| | **MF** | GO:0052689 | carboxylic ester hydrolase activity | 16·43 | 0·0020 | PLA2G1B/PNLIP/PNLIPRP1/PNLIPRP2 |
| | **BP** | GO:0071692 | protein localization to extracellular region | 10·48 | 0·0020 | CELA2A/GCG/PLA2G1B/PPY/SEL1L |

| | | | | | | |
|---|---|---|---|---|---|---|
| | **BP** | GO:0044241 | lipid digestion | 72·81 | 0·0064 | PNLIP/PNLIPRP2 |
| | **BP** | GO:0060193 | positive regulation of lipase activity | 72·81 | 0·0064 | PLA2G1B/PNLIP |
| | **MF** | GO:0004181 | metallocarboxypeptidase activity | 53·40 | 0·0114 | CPA2/CPB1 |
| | **BP** | GO:0060191 | regulation of lipase activity | 45·77 | 0·0145 | PLA2G1B/PNLIP |
| | **BP** | GO:0016042 | lipid catabolic process | 9·13 | 0·0145 | CLPS/PLA2G1B/PNLIP/PNLIPRP2 |
| | **BP** | GO:0002385 | mucosal immune response | 40·05 | 0·0179 | GP2/PLA2G1B |
| | **BP** | GO:0002251 | organ or tissue specific immune response | 36·41 | 0·0208 | GP2/PLA2G1B |
| | **MF** | GO:0008237 | metallopeptidase activity | 12·99 | 0·0220 | CPA2/CPB1/PRSS2 |
| | **MF** | GO:0004180 | carboxypeptidase activity | 34·08 | 0·0220 | CPA2/CPB1 |
| | **BP** | GO:0090279 | regulation of calcium ion import | 33·37 | 0·0221 | GCG/SPINK1 |
| | **BP** | GO:0090276 | regulation of peptide hormone secretion | 11·78 | 0·0261 | CELA2A/GCG/SPINK1 |

Abbreviations: GO – gene ontology, BP – biological process, MF – molecular function, CC – cellular component.

**Table S6. Top 30 KEGG pathways significantly enriched in each organ-specific proteomic clock.**

| Clock | Subcategory | ID | Description | Fold Enrichment | Adjusted *p*-value | Gene ID |
|---|---|---|---|---|---|---|
| **Adipose** | Endocrine system | hsa03320 | PPAR signaling pathway | 92·73 | 2·83e-05 | ADIPOQ/FABP4/PLIN1 |
| | Endocrine system | hsa04923 | Regulation of lipolysis in adipocytes | 79·64 | 0·0015 | FABP4/PLIN1 |
| | Endocrine system | hsa04920 | Adipocytokine signaling pathway | 67·12 | 0·0015 | ADIPOQ/LEP |
| | Signal transduction | hsa04152 | AMPK signaling pathway | 38·51 | 0·0035 | ADIPOQ/LEP |
| | Endocrine and metabolic disease | hsa04932 | Non-alcoholic fatty liver disease | 29·93 | 0·0046 | ADIPOQ/LEP |
| | NA | hsa04081 | Hormone signaling | 21·45 | 0·0073 | ADIPOQ/LEP |
| | Endocrine and metabolic disease | hsa04930 | Type II diabetes mellitus | 49·98 | 0·0397 | ADIPOQ |
| **Artery** | Cell motility | hsa04820 | Cytoskeleton in muscle cells | 22·10 | 0·0021 | BGN/ELN/THBS2 |
| **Brain** | Nervous system | hsa04721 | Synaptic vesicle cycle | 15·91 | 0·0011 | ATP6V1G2/DNM1/DNM3/SNAP25/SYT1 |
| | Infectious disease: bacterial | hsa05100 | Bacterial invasion of epithelial cells | 12·89 | 0·0102 | DNM1/DNM3/SEPTIN3/SEPTIN8 |
| **Heart** | Circulatory system | hsa04260 | Cardiac muscle contraction | 43·20 | 0·0125 | MYL4/TNNI3 |
| | Circulatory system | hsa04261 | Adrenergic signaling in cardiomyocytes | 24·41 | 0·0194 | MYL4/TNNI3 |
| | Cell motility | hsa04814 | Motor proteins | 19·08 | 0·0210 | MYL4/TNNI3 |
| | Cell motility | hsa04820 | Cytoskeleton in muscle cells | 16·20 | 0·0217 | MYL4/TNNI3 |
| **Immune** | Signaling molecules and interaction | hsa04060 | Cytokine-cytokine receptor interaction | 9·46 | 1·94e-09 | CCL5/CSF1R/CSF2RA/CSF3R/CXCL1/CXCL13/CXCL6/FASLG/IL18RAP/IL1R2/IL2RG/OSM/PF4/TNFRSF10C/TNFRSF1B |
| | Signaling molecules and interaction | hsa04061 | Viral protein interaction with cytokine and cytokine receptor | 18·79 | 4·30e-09 | CCL5/CSF1R/CXCL1/CXCL13/CXCL6/IL18RAP/IL2RG/PF4/TNFRSF10C/TNFRSF1B |
| | Development and regeneration | hsa04380 | Osteoclast differentiation | 10·51 | 2·17e-05 | CSF1R/FCGR2A/FCGR3B/LILRA5/LILRA6/LILRB2/NCF2/SIRPB1 |
| | Immune system | hsa04640 | Hematopoietic cell lineage | 13·16 | 2·17e-05 | CD7/CD8A/CSF1R/CSF2RA/CSF3R/IL1R2/ITGAM |
| | Immune system | hsa04613 | Neutrophil extracellular trap formation | 7·67 | 0·0002 | AZU1/FCGR2A/FCGR3B/ITGAM/MPO/NCF2/PADI4/SELPLG |
| | Transport and catabolism | hsa04145 | Phagosome | 7·09 | 0·0027 | CORO1A/FCGR2A/FCGR3B/ITGAM/MPO/NCF2 |
| | Signaling molecules and interaction | hsa04514 | Cell adhesion molecules | 7·05 | 0·0027 | CD8A/ICAM3/ITGAM/PTPRC/SELL/SELPLG |
| | Signal transduction | hsa04668 | TNF signaling pathway | 7·90 | 0·0051 | CCL5/CXCL1/CXCL6/MMP9/TNFRSF1B |
| | Cancer: overview | hsa05202 | Transcriptional misregulation in cancer | 5·61 | 0·0070 | CSF1R/GZMB/IL1R2/ITGAM/MMP9/MPO |
| | Immune system | hsa04650 | Natural killer cell mediated cytotoxicity | 7·01 | 0·0070 | FASLG/FCGR3B/GZMB/KIR2DL3/KIR2DS4 |
| | Infectious disease: parasitic | hsa05140 | Leishmaniasis | 9·52 | 0·0073 | FCGR2A/FCGR3B/ITGAM/NCF2 |

| | | | | | | |
|---|---|---|---|---|---|---|
| | Immune disease | hsa05340 | Primary immunodeficiency | 14·84 | 0·0088 | CD8A/IL2RG/PTPRC |
| | Immune system | hsa04662 | B cell receptor signaling pathway | 8·26 | 0·0105 | CD72/LILRA5/LILRA6/LILRB2 |
| | Immune disease | hsa05332 | Graft-versus-host disease | 12·53 | 0·0124 | FASLG/GZMB/KIR2DL3 |
| | Infectious disease: bacterial | hsa05150 | Staphylococcus aureus infection | 7·37 | 0·0138 | FCGR2A/FCGR3B/ITGAM/SELPLG |
| | Infectious disease: bacterial | hsa05152 | Tuberculosis | 5·16 | 0·0170 | CORO1A/FCGR2A/FCGR3B/ITGAM/TLR1 |
| | Immune system | hsa04062 | Chemokine signaling pathway | 4·87 | 0·0206 | CCL5/CXCL1/CXCL13/CXCL6/PF4 |
| | Cancer: specific types | hsa05221 | Acute myeloid leukemia | 8·29 | 0·0298 | CSF1R/ITGAM/MPO |
| | Cardiovascular disease | hsa05417 | Lipid and atherosclerosis | 4·35 | 0·0298 | CCL5/CXCL1/FASLG/MMP9/NCF2 |
| | Immune system | hsa04612 | Antigen processing and presentation | 6·96 | 0·0456 | CD8A/KIR2DL3/KIR2DS4 |
| **Intestine** | Digestive system | hsa04977 | Vitamin digestion and absorption | 40·16 | 0·0209 | APOA4/RBP2 |
| | Cancer: specific types | hsa05226 | Gastric cancer | 10·44 | 0·0209 | DH17/MUC2/REG4 |
| | Digestive system | hsa04975 | Fat digestion and absorption | 24·28 | 0·0209 | APOA4/FABP2 |
| | NA | hsa04081 | Hormone signaling | 7·15 | 0·0381 | GUCA2A/GUCY2C/MLN |
| | Endocrine system | hsa03320 | PPAR signaling pathway | 13·74 | 0·0381 | FABP2/FABP6 |
| **Kidney** | Lipid metabolism | hsa00100 | Steroid biosynthesis | 156·62 | 0·0256 | CYP24A1 |
| | Endocrine system | hsa04614 | Renin-angiotensin system | 136·19 | 0·0256 | REN |
| **Liver** | Immune system | hsa04610 | Complement and coagulation cascades | 50·85 | 8·64e-29 | C4BPB/C8B/C9/CFB/CFHR4/CFHR5/CPB2/F11/F12/F13B/F7/F9/FGA/KLKB1/MBL2/PLG/PROC/SERPINA1/SERPIND1/SERPINF2 |
| | Digestive system | hsa04979 | Cholesterol metabolism | 17·55 | 0·0027 | ANGPTL3/APOA2/APOC1/LPA |
| | Infectious disease: viral | hsa05171 | Coronavirus disease - COVID-19 | 5·64 | 0·0149 | C8B/C9/CFB/F13B/FGA/MBL2 |
| | Information processing in viruses | hsa03265 | Virion - Ebolavirus, Lyssavirus and Morbillivirus | 37·29 | 0·0231 | ASGR1/CLEC4M |
| **Pancreas** | Digestive system | hsa04972 | Pancreatic secretion | 62·58 | 3·31e-19 | AMY2A/CELA2A/CELA3A/CPA2/1360/1504/1506/5319/5406/5407/5408/5645 |
| | Digestive system | hsa04974 | Protein digestion and absorption | 36·85 | 3·94e-09 | CELA2A/CELA3A/CPA2/CPB1/CTRB1/CTRL/PRSS2 |
| | Digestive system | hsa04975 | Fat digestion and absorption | 64·27 | 7·81e-08 | CLPS/PLA2G1B/PNLIP/PNLIPRP1/PNLIPRP2 |
| | Lipid metabolism | hsa00561 | Glycerolipid metabolism | 25·51 | 0·0013 | PNLIP/PNLIPRP1/PNLIPRP2 |

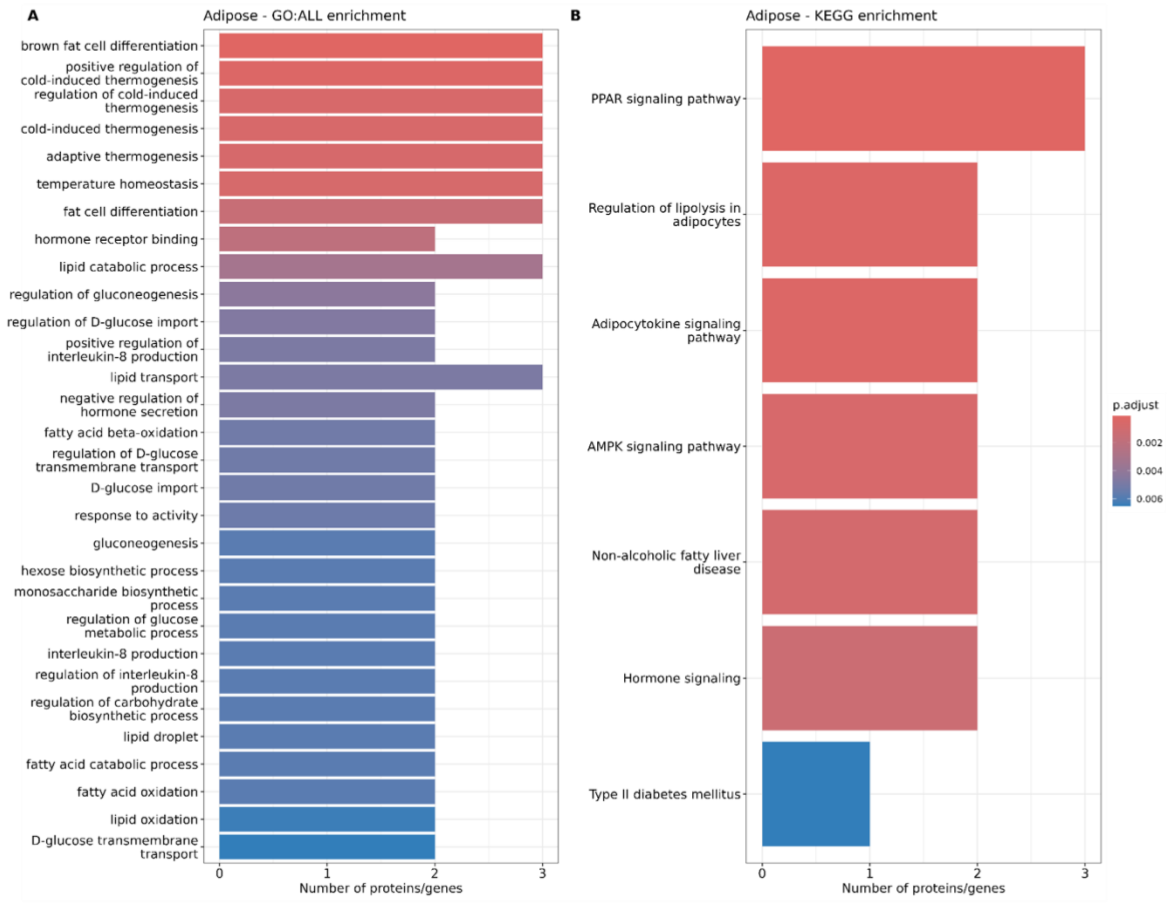


**Figure S2. Top 30 GO categories and KEGG pathways significantly enriched in the adipose clock.**

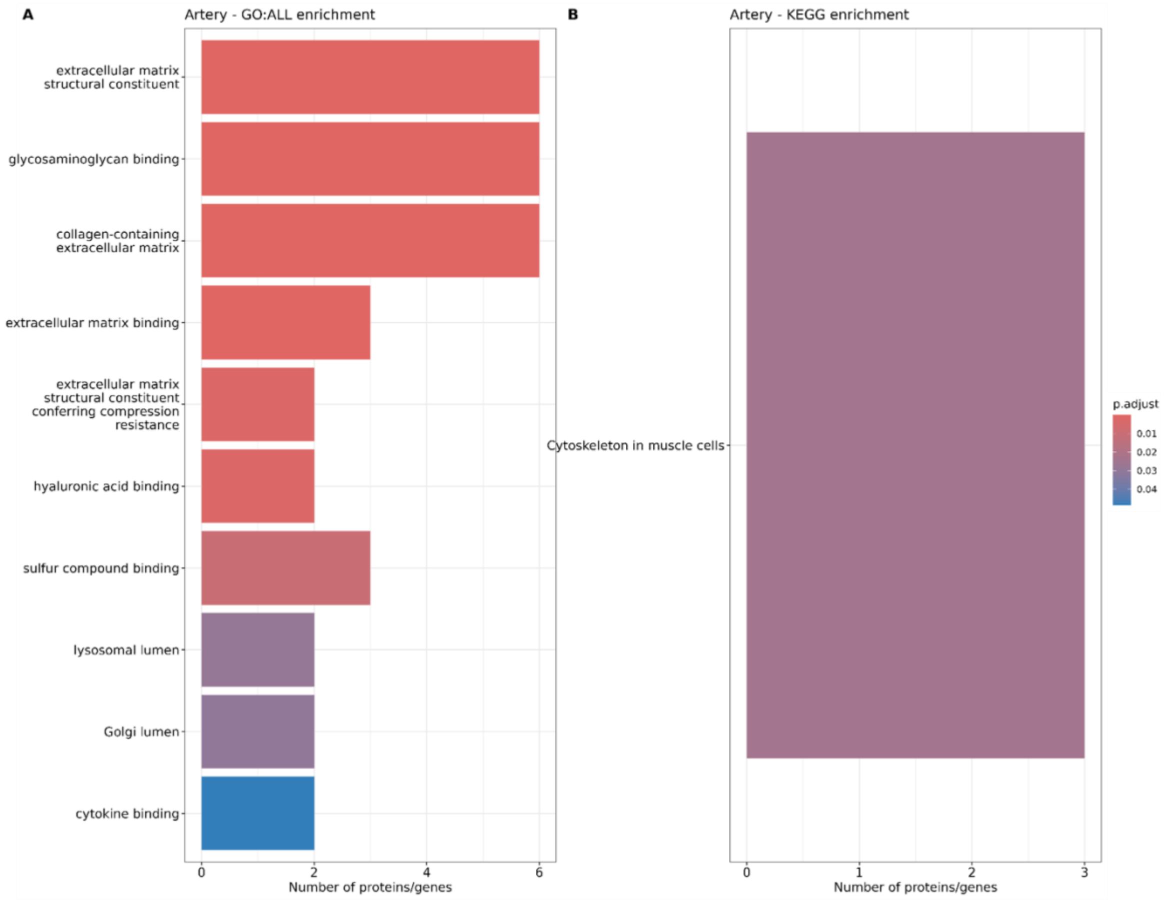


**Figure S3. Top 30 GO categories and KEGG pathways significantly enriched in the artery clock.**

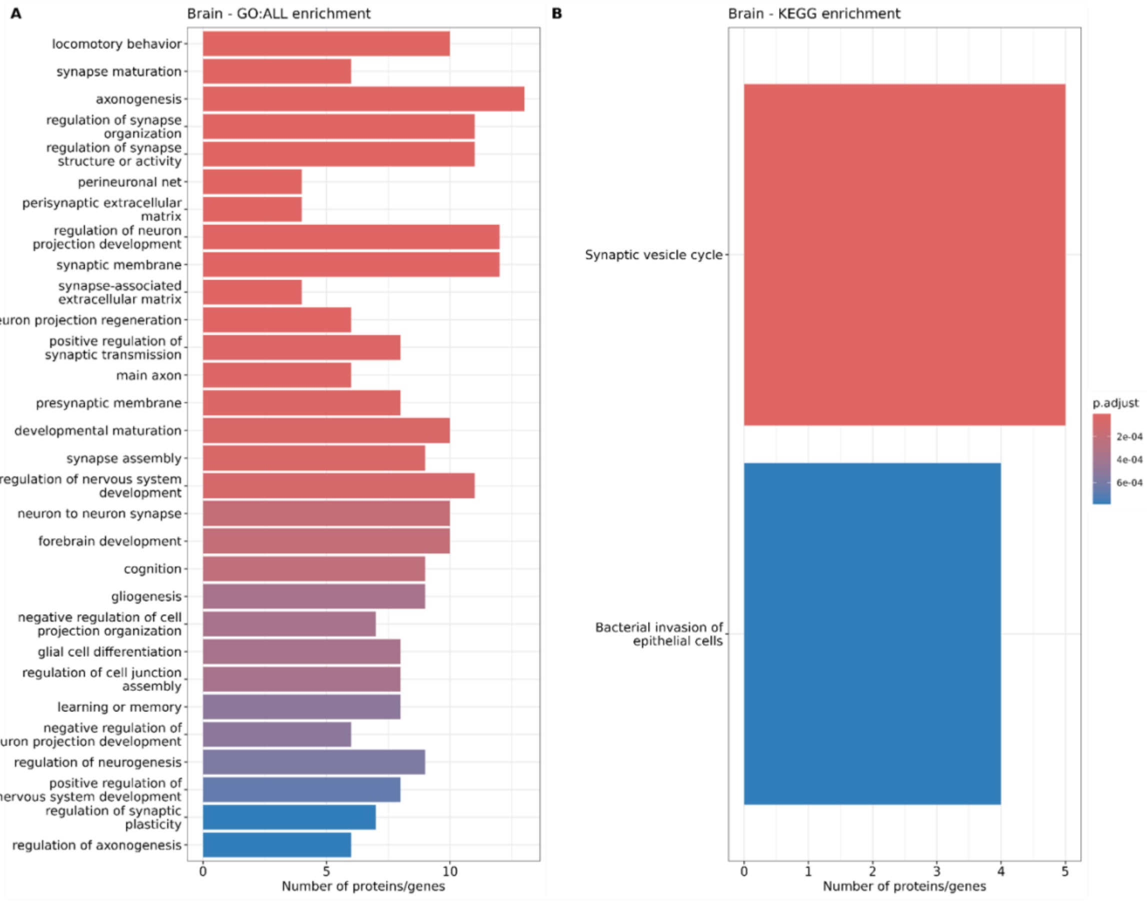


**Figure S4. Top 30 GO categories and KEGG pathways significantly enriched in the brain clock.**

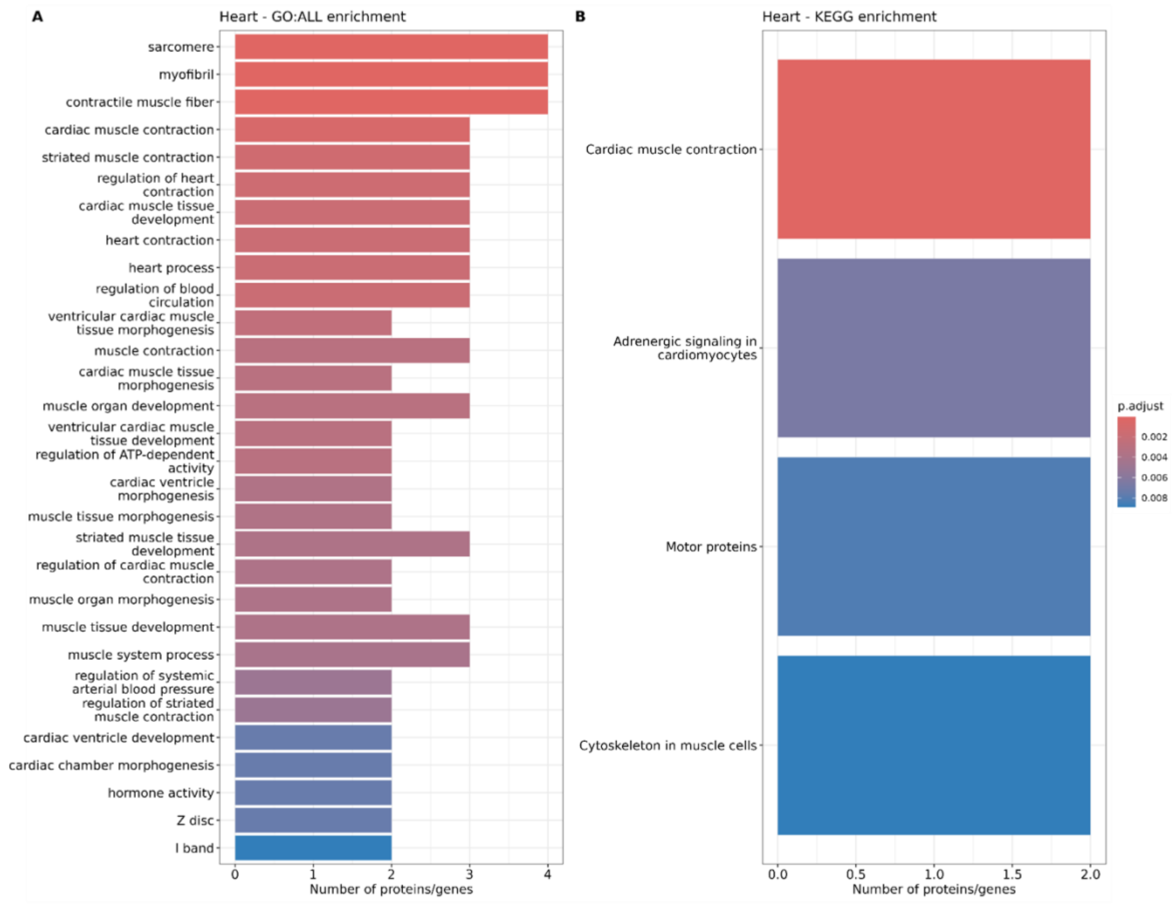


**Figure S5. Top 30 GO categories and KEGG pathways significantly enriched in the heart clock.**

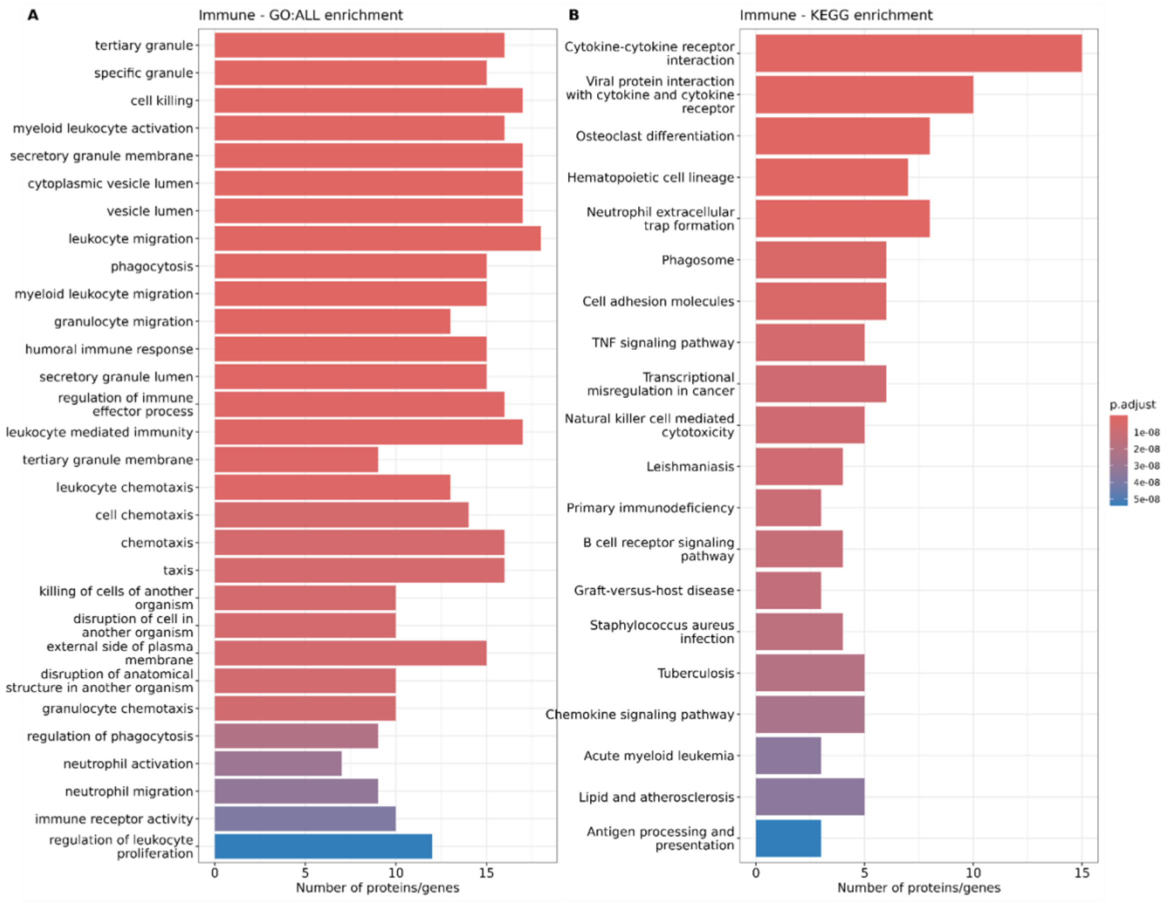


**Figure S6. Top 30 GO categories and KEGG pathways significantly enriched in the immune clock.**

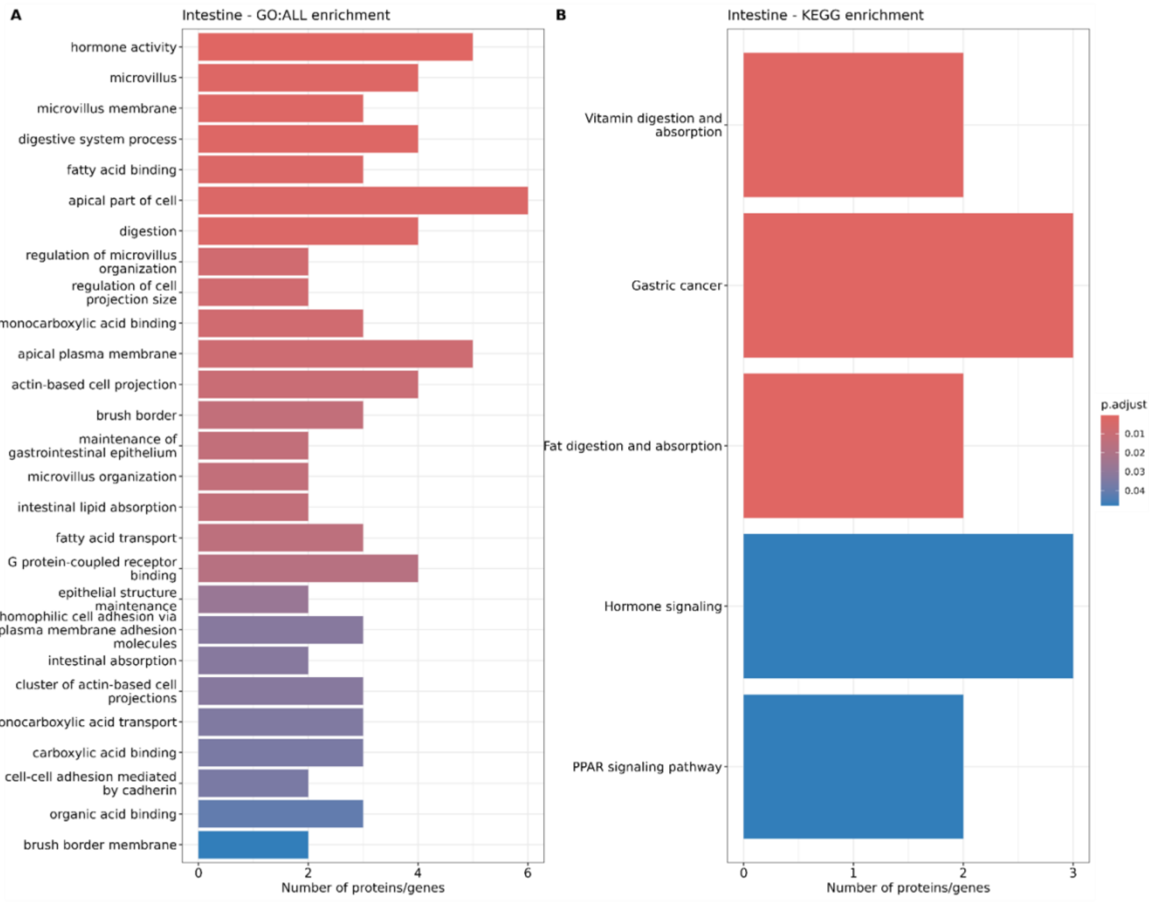


**Figure S7. Top 30 GO categories and KEGG pathways significantly enriched in the intestine clock.**

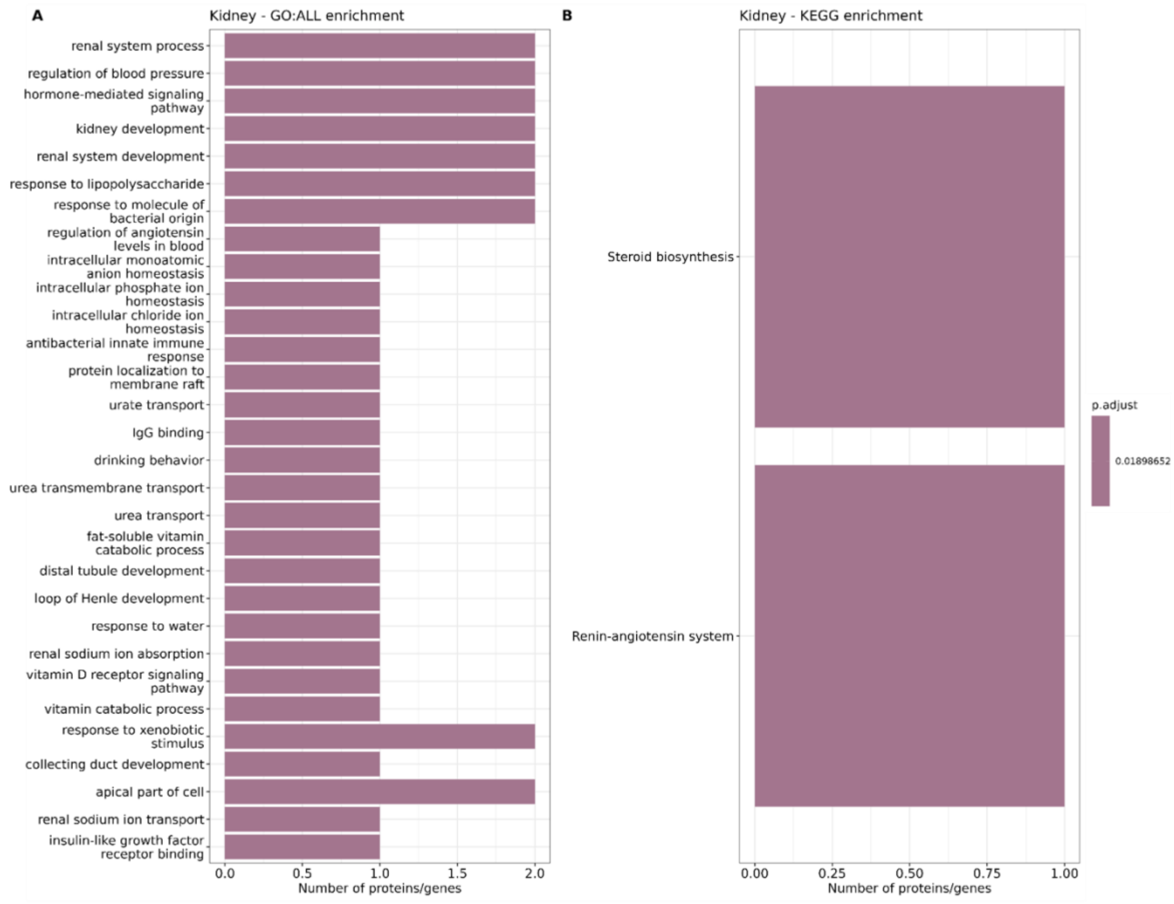


**Figure S8. Top 30 GO categories and KEGG pathways significantly enriched in the kidney clock.**

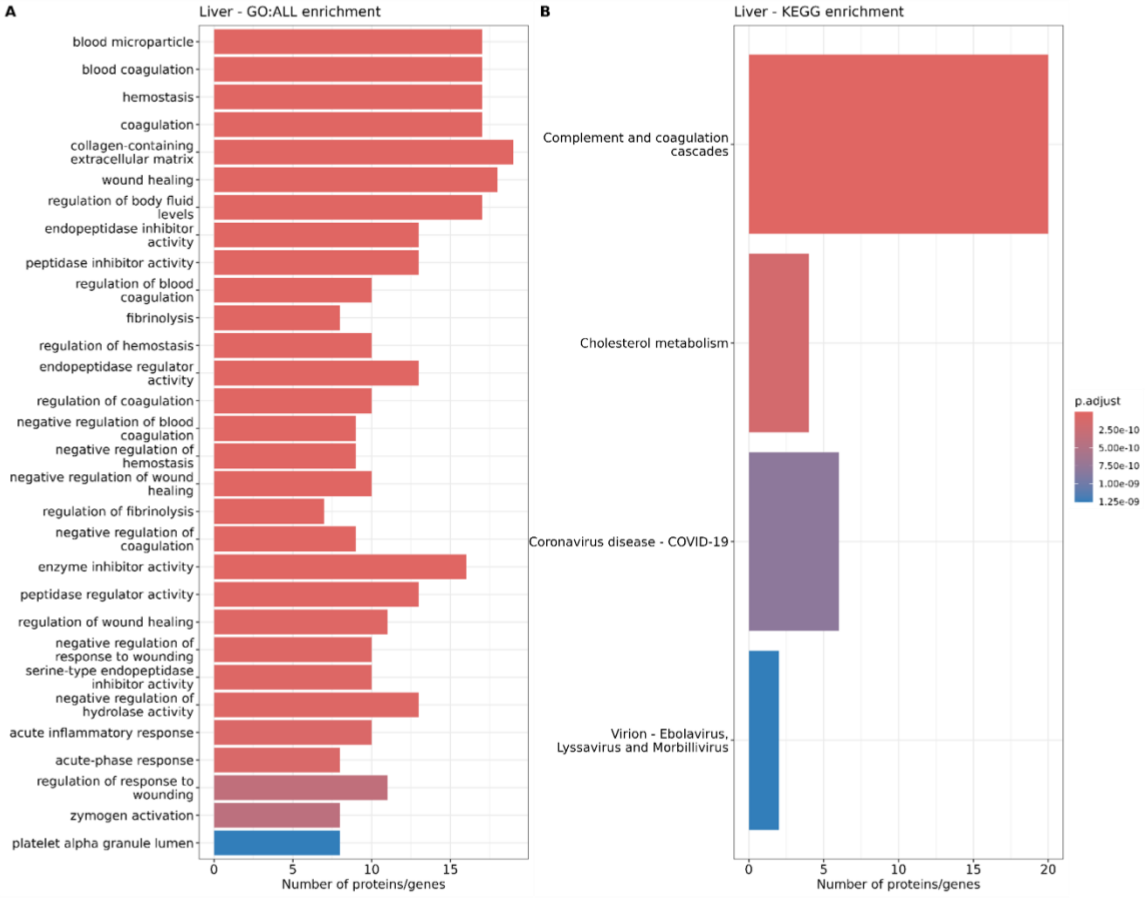


**Figure S9. Top 30 GO categories and KEGG pathways significantly enriched in the liver clock.**

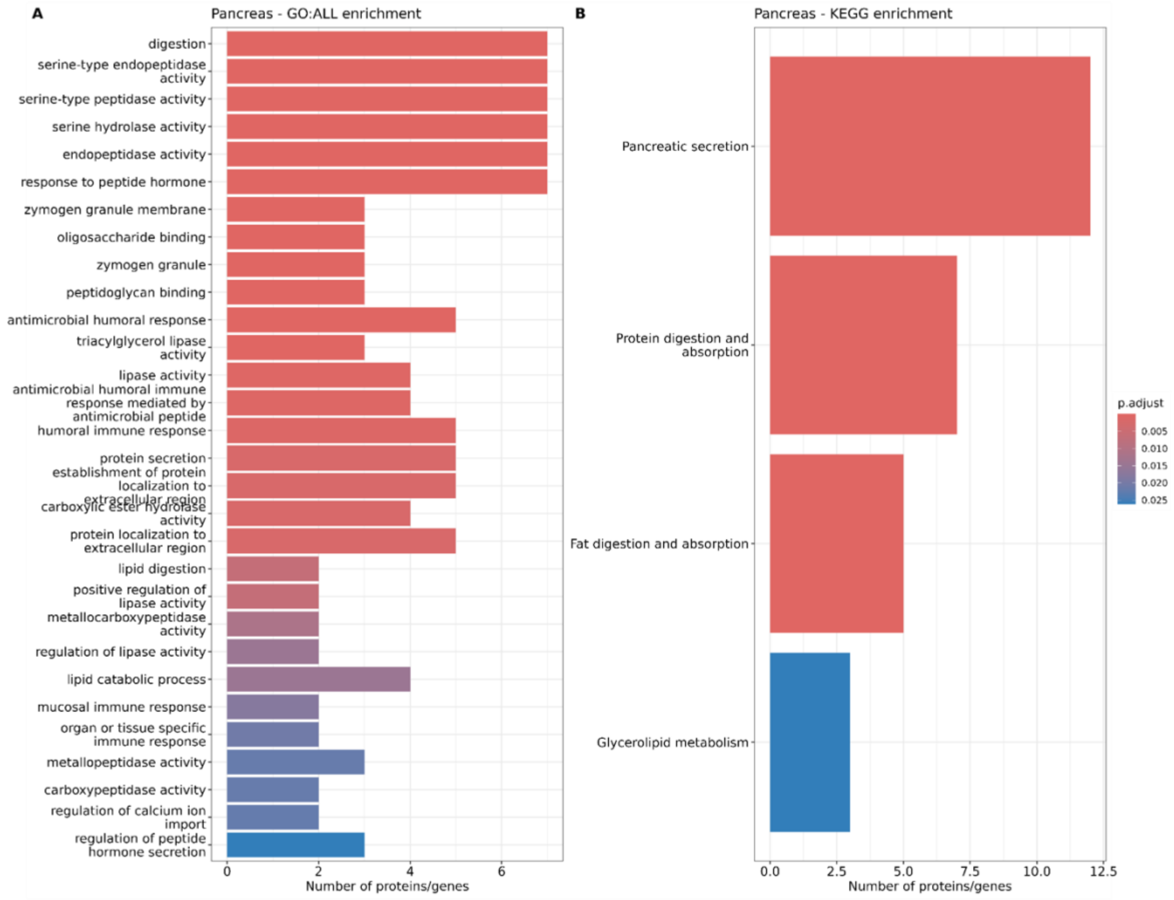


**Figure S10. Top 30 GO categories and KEGG pathways significantly enriched in the pancreas clock.**

**Table S7. Variance inflation factors for each predictor (clock) considered for multivariate analysis.**

| | GCS | TICS-m3 | RT | p-tau217 | GFAP | NfL | Level of collinearity |
|---|---|---|---|---|---|---|---|
| **Heart_2ndGen** | 1·37 | 1·38 | 1·47 | 1·37 | 1·40 | 1·40 | Low |
| **Adipose_2ndGen** | 1·47 | 1·47 | 1·36 | 1·46 | 1·52 | 1·52 | |
| **Kidney_2ndGen** | 1·52 | 1·56 | 1·56 | 1·53 | 1·55 | 1·55 | |
| **Liver_2ndGen** | 1·68 | 1·70 | 1·66 | 1·70 | 1·69 | 1·69 | |
| **Pancreas_2ndGen** | 1·74 | 1·78 | 1·66 | 1·76 | 1·76 | 1·76 | |
| **Intestine_2ndGen** | 1·79 | 1·84 | 1·84 | 1·83 | 1·81 | 1·81 | |
| **Artery_2ndGen** | 1·98 | 2·03 | 2·21 | 1·99 | 1·99 | 1·99 | |
| **Immune_2ndGen** | 2·13 | 2·15 | 2·22 | 2·16 | 2·18 | 2·18 | |
| **DunedinPACE** | 3·02 | 3·06 | 2·93 | 3·06 | 3·03 | 3·03 | |
| **Brain_2ndGen** | 3·16 | 3·12 | 2·98 | 3·16 | 3·15 | 3·15 | Moderate |
| **1 − HPS** | 6·14 | 5·96 | 6·44 | 6·04 | 6·09 | 6·09 | |
| **PhenoAge** | 6·12 | 6·00 | 5·30 | 6·16 | 6·25 | 6·25 | |
| **Horvath** | 7·83 | 7·45 | 7·45 | 7·61 | 7·60 | 7·60 | |
| **GrimAge** | 8·37 | 8·28 | 8·09 | 8·20 | 8·18 | 8·18 | |
| **GrimAge2** | 8·79 | 8·99 | 8·37 | 8·45 | 8·39 | 8·39 | |
| **PAC** | 8·97 | 8·70 | 9·19 | 8·82 | 8·83 | 8·83 | |
| **Hannum** | 9·41 | 9·04 | 8·50 | 9·12 | 9·26 | 9·26 | |
| **Conventional_2ndGen** | 12·18 | 12·33 | 12·81 | 12·18 | 12·14 | 12·14 | High |

Abbreviations: GCS – Global Cognitive Score; TICS-m3 – Telephone Interview for Cognitive Status modified version 3; RT – Reaction Time; p-tau217 – phosphorylated tau 217; GFAP – glial fibrillary acidic protein; NfL – neurofilament light chain; HPS – Healthspan Proteomic Score; PAC – Proteomic Aging Clock.

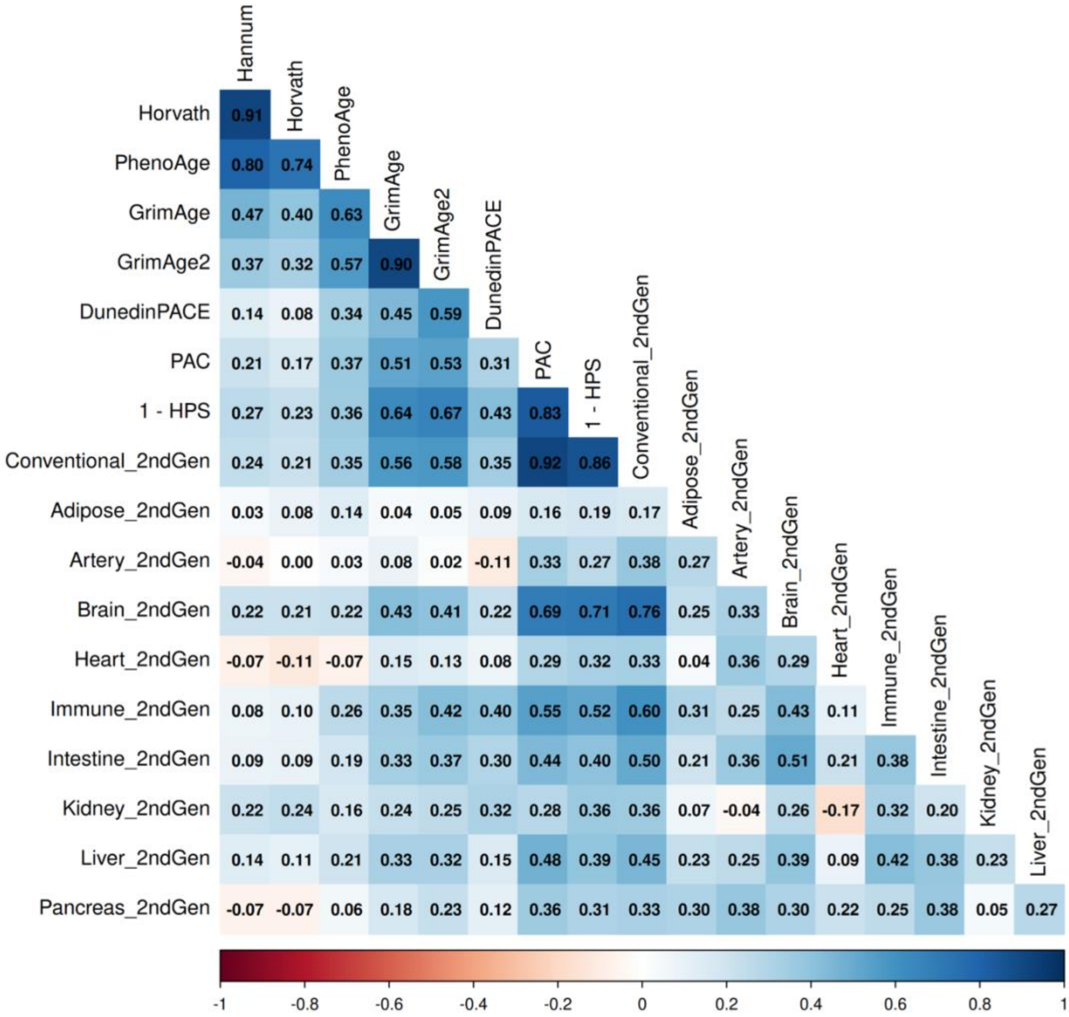


**Figure S11. Pearson's correlations between each proteomic and epigenetic aging clock.**

**Table S8. Decrease in pseudo-$R^2$ in multivariate submodels.**

| Dropped predictor | GCS | TICS-m3 | RT | p-tau217 | GFAP | NfL |
|---|---|---|---|---|---|---|
| **PAC** | 0·0045 | 0·0037 | 0·0219 | 0·0105 | 0·0076 | 0·0061 |
| **1 − HPS** | 0·0054 | 0·0027 | -0·0075 | 0·0002 | 0·0088 | 0·0115 |
| **Adipose_2ndGen** | 0·0049 | 0·0038 | 0·0382 | 0·0085 | 0·0221 | 0·0029 |
| **Artery_2ndGen** | 0·0135 | -2·6973e-05 | 0·0034 | 0·0035 | 0·0202 | 0·0476 |
| **Brain_2ndGen** | 0·0067 | 0·0059 | -0·0007 | 0·0004 | 0·0025 | 0·0061 |
| **Heart_2ndGen** | 0·0002 | 0·0020 | -0·0114 | 0·0010 | 0·0034 | 0·0020 |
| **Immune_2ndGen** | 0·0056 | 0·0184 | 0·0105 | 2·1472e-05 | 0·0004 | -4·8487e-05 |
| **Intestine_2ndGen** | 0·0004 | 0·0134 | 0·0316 | 0·0034 | 0·0069 | 0·0054 |
| **Kidney_2ndGen** | 0·0208 | 0·0414 | 0·0031 | 0·0180 | 5·7122e-05 | 0·0094 |
| **Liver_2ndGen** | 0·0041 | 0·0096 | 0·0163 | -3·9649e-05 | -6·5647e-05 | 0·0010 |
| **Pancreas_2ndGen** | 0·0370 | 0·0100 | 0·0450 | 0·0158 | 0·0016 | 0·0080 |

Higher values indicate higher importance of the predictor in the model.
Abbreviations: GCS – Global Cognitive Score; TICS-m3 – Telephone Interview for Cognitive Status modified version 3; RT – Reaction Time; p-tau217 – phosphorylated tau 217; GFAP – glial fibrillary acidic protein; NfL – neurofilament light chain; PAC – Proteomic Aging Clock; HPS – Healthspan Proteomic Score.
